# Functional Connectivity of the Neonatal Cerebellum is Impacted by Sex and Polygenic Liability for Autism

**DOI:** 10.64898/2026.04.17.26351076

**Authors:** Lauren Wagner, Emily Chiem, Janelle Liu, Leanna M Hernandez

## Abstract

The cerebellum rapidly integrates with cerebral networks during infancy and shows consistent structural and functional alterations in Autism Spectrum Disorder (ASD), suggesting that early cerebellar development may be consequential for later behavioral and psychiatric outcomes. Yet, little is known about the effect of ASD genetic liability on cerebello-cerebral functional connectivity in infancy or whether effects may differ by biological sex. Here, we leveraged neonatal functional magnetic resonance imaging, genetic, and behavioral follow-up data from the Developing Human Connectome Project (dHCP) to examine the relationship between ASD polygenic scores (PGS) and functional connectivity of cerebellar regions associated with sensorimotor and social-cognitive functions in 198 term-born neonates (mean age: 9.7 days). We report widespread sex differences in neonatal cerebello-cerebral connectivity that are regionally specific across cerebellar subdivisions. Across the full sample, elevated ASD PGS predicted alterations in cerebello-cerebral connectivity, with hemisphere-dependent differences in sensorimotor cerebellar connectivity with temporal cortex, and hyperconnectivity between the right social-cognitive seed and posterior cingulate. Notably, elevated ASD PGS predicted opposing patterns of cerebello-cerebral connectivity in males and females, including male hyperconnectivity between the right sensorimotor cerebellum and default mode areas, and female hyperconnectivity between the right social-cognitive seed and sensorimotor cortex. Connectivity associated with elevated ASD PGS showed nominal, sex-specific associations with 18-month language ability, attention problems, and emotional reactivity. Our findings show that ASD PGS influences the functional configuration of the cerebellum at birth and suggest that underlying cerebellar connectivity profiles associated with ASD may partially underlie distinct behavioral presentations in males and females.

## Introduction

Autism Spectrum Disorder (ASD) is a highly heritable neurodevelopmental condition (1) that affects roughly 1 in 31 individuals (2) and is approximately four times more prevalent in males than females. ASD is characterized by difficulties in social communication and the presence of restricted, repetitive behaviors, and its effects on early brain development may be detectable even before birth (3). Population-based studies suggest that approximately 80% of the variation in ASD-related traits is attributable to genetic factors (4). A substantial portion of genetic liability for ASD is thought to arise from the additive effects of many common risk variants across the genome (5–8) which, individually, confer small effect sizes on the likelihood for developing ASD but, in aggregate, may substantially impact liability for atypical development and ultimate diagnosis.

ASD-associated variants identified though genome wide association studies (GWAS) are enriched in genes involved in neuronal function and brain development, including synaptic transmission and plasticity (9), as well as neuronal differentiation, axonal and dendritic development, and corticogenesis (10). This genetic architecture aligns with the highly developmental nature of ASD, which has a global mean diagnosis age of 5 years of age (11) and can be reliably diagnosed as early as 18 months (12). Consistent with this, prior work has linked common ASD-associated risk variants to differences in brain structure and function in children and adults, including cerebellar and brainstem volume (13), functional connectivity of the salience network (14), cortical thickness and white matter connectivity (15), and neurite density (16). However, because the neurodevelopmental processes implicated by ASD GWAS are most active during early brain development, it is critical to examine how common genetic liability for ASD shapes the brain during infancy, before the onset of overt behavioral symptoms. To date, only one study has investigated associations between ASD genetic liability and brain function during infancy (17), highlighting a substantial gap in our understanding of the earliest neural consequences of genetic risk.

The relationship between genetic liability for ASD and infant cerebellar function remains, to our knowledge, wholly unstudied. Although only one tenth the mass of the cerebrum (18), the cerebellum contains more neurons than the rest of the brain combined (19). Traditionally known for its role in motor-related learning, error detection, and coordination, the “little brain” has more recently been recognized for its role in non-motor behaviors such as cognition, language, attention, emotional regulation, social communication, and perceptual processing (see (20–23) for reviews). Because the foundations of these functions are established during infancy, studying cerebellar function soon after birth is likely to provide insights into early differences in behavioral development.

Importantly, the cerebellum consistently shows alterations in children and adults with ASD at the functional, structural, and cellular levels (see (24,25) for reviews). For example, the cerebellum shows atypical activity during motor tasks in ASD (26,27), and reductions in cerebellar gray matter volume may relate to ASD-associated language delay (28). At the cellular level, loss of inhibitory Purkinje neurons is commonly reported in ASD postmortem cortex (25,29), supporting the notion that the cerebellum’s regulatory effects on the cortex may be disrupted in autism (30). Notably, the cerebellum has also been implicated in ASD-associated symptoms across development, including sensory sensitivity (31,32), social-communication deficits (33), and broader autism symptoms (34,35). Individuals with autism also show reliable functional underconnectivity between the cerebellar crus I/II – typically associated with higher-order cognition and language – and prefrontal, parietal, and frontotemporal association areas (32,34–36), whereas these cerebellar cognitive lobules are overconnected with sensorimotor cortex (32). Together, these findings point to a model in which disrupted cerebellar regulation of distributed cortical networks contributes to both sensorimotor and higher-order cognitive features of ASD.

Cerebello-cortical networks are established early in life, shifting from greater intracerebellar connectivity in mid-infancy (37) to greater cerebello-cerebral connectivity in adulthood (38). Despite recent work indicating that the cerebellum changes most dynamically during the first year of life (39), underscoring its potentially foundational role in broader neurodevelopment, few studies have investigated potential links between cerebellar function and autism in infancy. Existing work suggests that atypical cerebellar function in infants with an older ASD-diagnosed sibling (who are at high likelihood, HL, for autism) may be related to differences in language processing (40) and developmental language outcomes (41). The recruitment of HL infants has spawned a substantial literature over several decades (42,43), using familial risk as a proxy for underlying genetic liability for ASD. However, HL infants are not a uniform group: while approximately 20% will later meet diagnostic criteria for ASD (44,45), others exhibit subclinical developmental differences (46), and many develop typically. Advances in GWAS, combined with large-scale infant neuroimaging datasets with genotype data, now make it possible to extend the family-based “Baby Sibs” paradigm by directly quantifying polygenic liability for ASD at the population level and to test for associations with brain-based phenotypes. Despite these advances, studies investigating how common genetic variation impacts the developing brain remain rare, and little is known about how common genetic liability for ASD shapes cerebellar functional organization in early life.

To address this gap, we integrated MRI, genetic, and behavioral data from the Developing Human Connectome Project (dHCP; (47)) to examine how polygenic liability for autism influences the functional configuration of the cerebellum at birth, as well as 18-month behavioral outcomes. We focused on common genetic variation indexed by ASD polygenic scores and assessed their associations with cerebellar subdivisions linked to primary sensorimotor and higher-order cognitive functions. Given well-established sex differences in ASD prevalence, we further tested whether these associations differed by biological sex.

## Methods

### Participants

Participants were recruited and enrolled at St Thomas’ Hospital, London as part of the Developing Human Connectome Project (dHCP; (47)), a large-scale study of fetal and neonatal brain development. All data were obtained from the publicly available dHCP data release through the National Institute of Mental Health National Data Archive (https://nda.nih.gov/). Data collection was approved by the UK National Research Ethics Authority (14/LO/1169) and informed written consent was given by participants’ parents or primary caregivers.

The dHCP protocol collected MRI scans between 0-138 days (mean: 19; median: 12 days) after birth and excluded infants with contraindication to MRI (e.g., metallic implants, an inability to tolerate scanning due to extreme prematurity and/or serious medical complications, or a language barrier that prevented communication with caregivers). Infants born at term, defined as 37–44 weeks gestational age (GA), were included in the present study. Our final sample consisted of 198 infants with quality-checked fMRI and genetic data (mean postnatal age at scan (PNA): 9.73 days, mean GA at birth: 40.18 weeks).

### Behavioral Assessments

Behavioral follow-up assessments measured multiple cognitive and developmental milestones at 18 months. Given the cerebellum’s consistent implication in ASD pathology and its role in emerging motor, cognitive, attentional, and language development, we focused on ASD symptoms as assessed by the Quantitative Checklist for Autism in Toddlers (Q-CHAT; (48)); the cognitive, motor, and language subscales of the Bayley Scales of Infant and Toddler Development, Third Edition (BSITD-III; (49)); and the Child Behavioral Checklist (CBCL; (50)) subscales of sleep, attention problems, and emotional reactivity. Mean scores and sex differences are shown in Table 1.

**Table 1.**
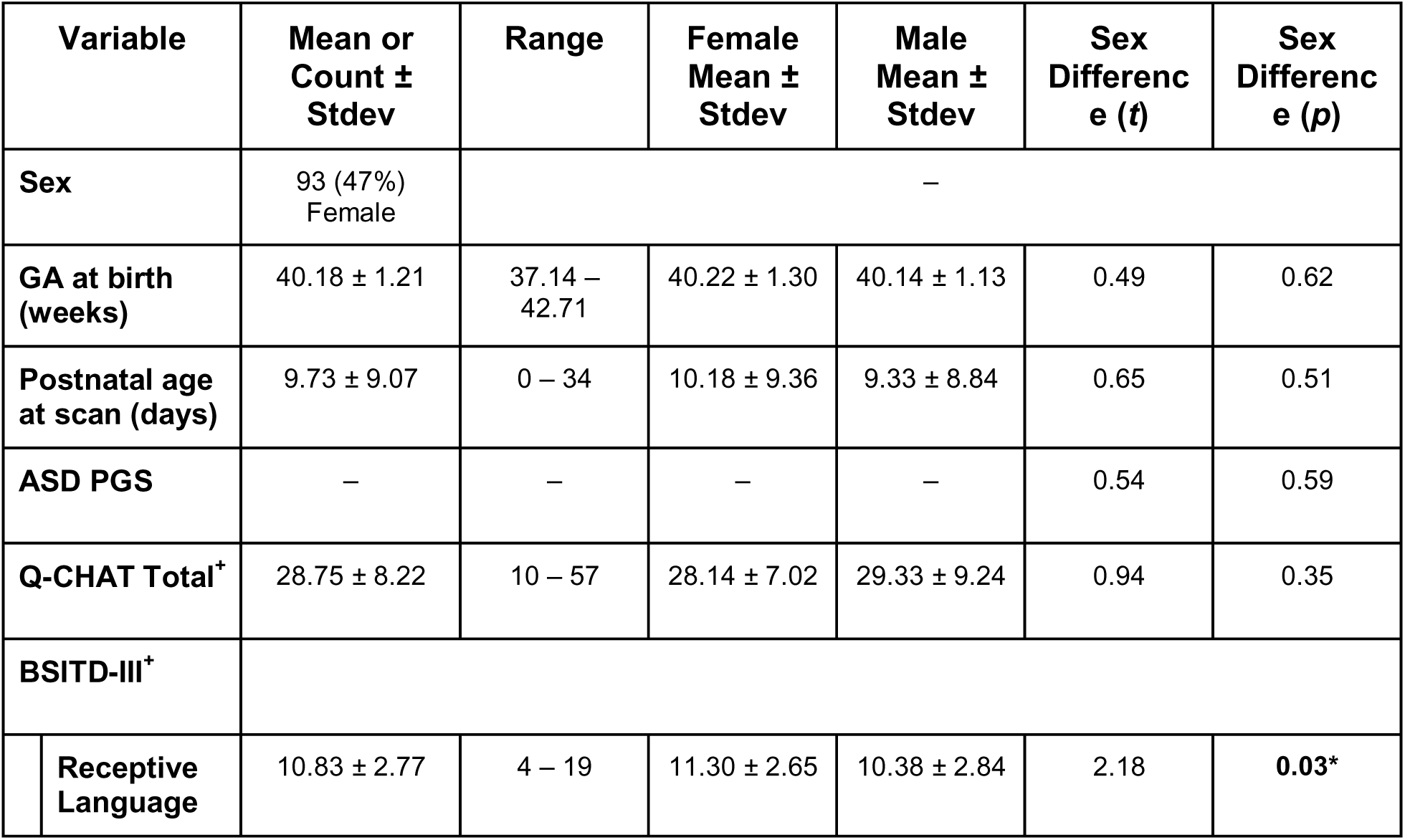
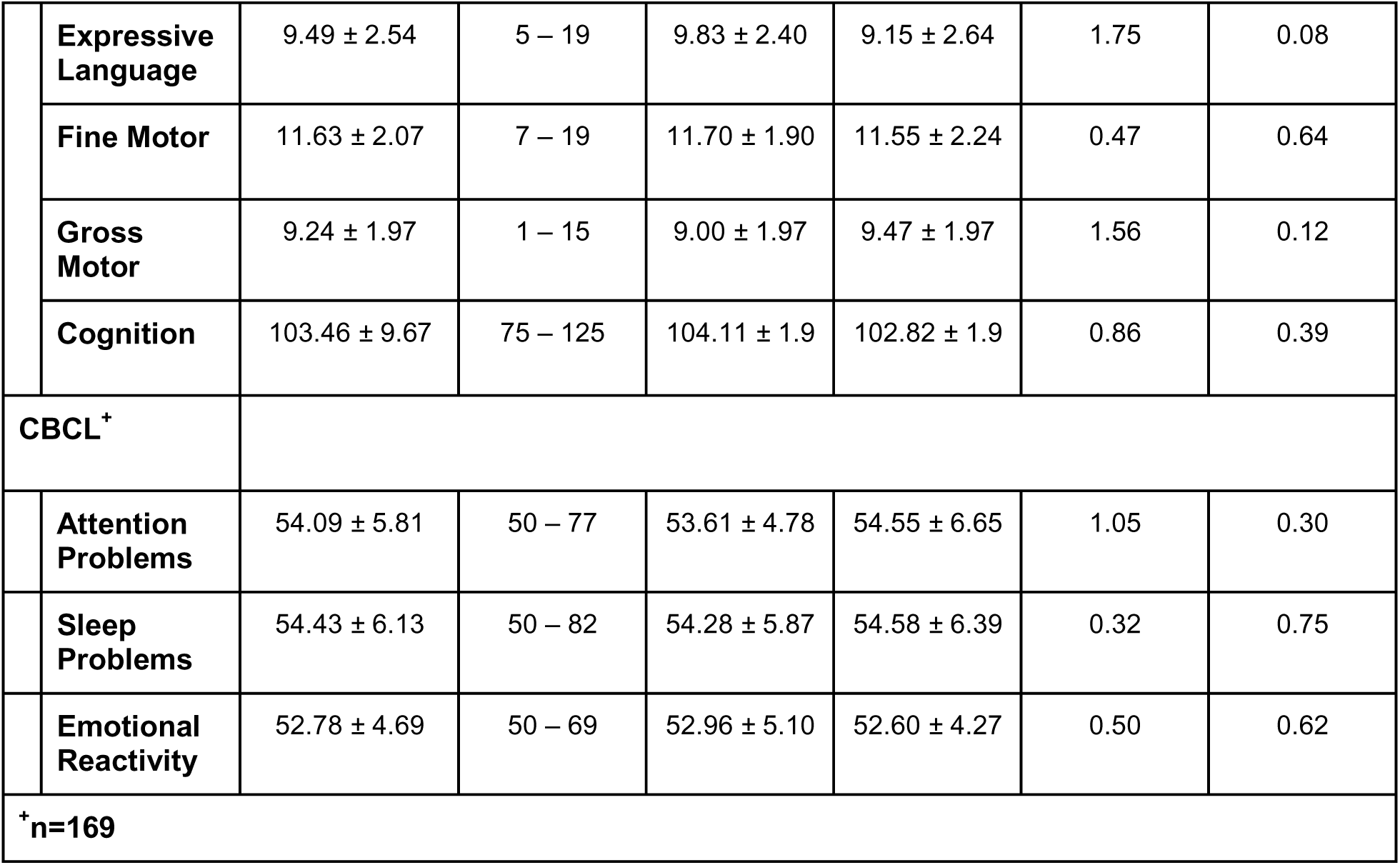
Participant characteristics and behavioral outcome variables for the whole sample (N=198) and the subset with behavioral outcome data (N=169). Sex differences were assessed using a two-tailed independent samples t-test for normally distributed continuous measures. Stdev = Standard Deviation; GA = Gestational Age; ASD PGS = Autism Polygenic Score; Q-CHAT = Quantitative Checklist for Autism in Toddlers; BSITD-III = Bayley Scales of Infant and Toddler Development, Third Edition.

| Variable | Mean or Count ± Stdev | Range | Female Mean ± Stdev | Male Mean ± Stdev | Sex Difference (*t*) | Sex Difference (*p*) |
| --- | --- | --- | --- | --- | --- | --- |
| Sex | 93 (47%)<br>Female | – |  |  |  |  |
| GA at birth (weeks) | 40.18 ± 1.21 | 37.14 – 42.71 | 40.22 ± 1.30 | 40.14 ± 1.13 | 0.49 | 0.62 |
| Postnatal age at scan (days) | 9.73 ± 9.07 | 0 – 34 | 10.18 ± 9.36 | 9.33 ± 8.84 | 0.65 | 0.51 |
| ASD PGS | – | – | – | – | 0.54 | 0.59 |
| Q-CHAT Total<sup>+</sup> | 28.75 ± 8.22 | 10 – 57 | 28.14 ± 7.02 | 29.33 ± 9.24 | 0.94 | 0.35 |
| BSITD-III<sup>+</sup> |  |  |  |  |  |  |
| Receptive Language | 10.83 ± 2.77 | 4 – 19 | 11.30 ± 2.65 | 10.38 ± 2.84 | 2.18 | **0.03\*** |

**Table 1.** Participant characteristics and behavioral outcome variables for the whole sample (N=198) and the subset with behavioral outcome data (N=169).
|  |  |  |  |  |  |  |  |
| --- | --- | --- | --- | --- | --- | --- | --- |
|  | **Expressive Language** | 9.49 ± 2.54 | 5 – 19 | 9.83 ± 2.40 | 9.15 ± 2.64 | 1.75 | 0.08 |
|  | **Fine Motor** | 11.63 ± 2.07 | 7 – 19 | 11.70 ± 1.90 | 11.55 ± 2.24 | 0.47 | 0.64 |
|  | **Gross Motor** | 9.24 ± 1.97 | 1 – 15 | 9.00 ± 1.97 | 9.47 ± 1.97 | 1.56 | 0.12 |
|  | **Cognition** | 103.46 ± 9.67 | 75 – 125 | 104.11 ± 1.9 | 102.82 ± 1.9 | 0.86 | 0.39 |
| **CBCL<sup>+</sup>** |  |  |  |  |  |  |  |
|  | **Attention Problems** | 54.09 ± 5.81 | 50 – 77 | 53.61 ± 4.78 | 54.55 ± 6.65 | 1.05 | 0.30 |
|  | **Sleep Problems** | 54.43 ± 6.13 | 50 – 82 | 54.28 ± 5.87 | 54.58 ± 6.39 | 0.32 | 0.75 |
|  | **Emotional Reactivity** | 52.78 ± 4.69 | 50 – 69 | 52.96 ± 5.10 | 52.60 ± 4.27 | 0.50 | 0.62 |
| **<sup>+</sup>n=169** |  |  |  |  |  |  |  |

### Genetic Processing & Polygenic Score Calculation

SNPs were filtered for minor allele frequency <0.01, missingness per individual >0.01, missingness per marker >0.01, and Hardy-Weinberg equilibrium (p<10^-6^) using PLINK v2.0. Genotype data were then imputed to the TOPMed reference panel using the TOPMed Imputation Server (imputationserver.sph.umich.edu). Using data from the 1000 Genomes Project (51) as reference, a K-nearest neighbors clustering algorithm was applied to assign categorical genetic ancestry, and principal components reflecting genetic ancestry were calculated in PLINK. For participants assigned to European ancestry, genotype data was used to calculate polygenic scores (PGS) for ASD, providing an aggregate score that quantifies the cumulative common genetic risk for ASD across the genome. PGS were calculated using summary statistics from the largest published genome-wide association study (GWAS) of ASD (7), composed of 18,381 ASD cases and 27,969 controls. ASD PGS were computed using PRS-CS (52) using an ancestry-matched LD reference panel from the 1000 Genomes Project and the recommended global shrinkage parameter for highly polygenic traits (phi=1e-2). All subsequent analyses involving the PGS included the first five principal ancestry components as covariates to control for residual confounding due to population stratification.

### Image Acquisition

Resting-state and T2-weighted structural magnetic resonance images (MRIs) were collected on a 3T Philips Achieva scanner at King’s College London using a 32-channel neonatal head coil. Scans were conducted during natural sleep after infant participants were fed and swaddled. Hearing protection was provided in the form of dental putty, protective earmuffs, and an acoustic hood. Participants were monitored for pulse oximetry, respiration, and body temperature. Scan sequences were optimized for the properties of the neonatal brain (see (47) and the supplemental methods for more information).

### fMRI Preprocessing

DHCP data were downloaded from the National Institute of Mental Health National Data Archive (NDA). MRI scans underwent minimal preprocessing as described in (53), which included volume-to-volume and slice-to-volume motion correction, spatial normalization to the 40-week GA dHCP template, susceptibility distortion correction, denoising using ICA-FIX, and automated quality control. We then applied additional processing steps using FMRIB’s Software Library (FSL; (54)) in preparation for functional connectivity analyses, including: spatial smoothing (3mm kernel), bandpass filtering (0.01<*t*<0.1 Hz), and the regression of white matter, cerebrospinal fluid, and global signal.

### fMRI Data Analysis

We employed a seed-based functional connectivity analysis to examine voxelwise connectivity between cerebellar regions of interest (ROIs) and the rest of the brain. Because functional specificity in cerebellar subregions is relatively immature at birth (39), we combined functionally similar regions into SensoriMotor (SM) and SocialCognitive (SC) ROIs. Informed by lesion studies and task batteries interrogating cerebellar function (55–57), as well as prior functional connectivity studies of cerebellar subregions (31,58), the SM ROI consisted of Lobules I-IV, V, VIIIa, and VIIIb, while the SC ROI consisted of Crus I/II (Figure S1). The average timeseries was extracted from each ROI at the single-subject level and correlated with that of every voxel in the cerebrum. Correlation maps were transformed into *z*-statistic maps using Fisher’s *r*-to-*z* transformation. Group-level analyses were then performed in FSL using FMRIB’s Local Analysis of Mixed Effects (FLAME 1). Group-average maps of connectivity were generated for each cerebellar ROI, controlling for sex, GA at birth, and postnatal age at the time of scan (PNA). Sex differences in the connectivity of each ROI were also examined, controlling for GA at birth and PNA at scan. Next, we examined the relationship between genetic liability for ASD and cerebellar functional connectivity by using the previously computed ASD PGS as a predictor in the whole-brain regression analysis, controlling for sex, GA at birth, PNA, and the first five principal ancestry components. Finally, we examined the interaction between ASD PGS and biological sex, controlling for age and ancestry.

Across all voxelwise results, regions whose peaks exceeded a significance threshold of *Z*>3.1 are reported (Tables S1 and S2). Connectivity maps are displayed at multiple thresholds (*Z*>2.3, *Z*>2.7; equivalent to approximately *p*<0.01 and *p*<0.003, respectively) to illustrate the spatial extent and strength of the observed effects, correcting for multiple comparisons at the cluster level (*p*<0.05). Because analyses of sex differences were especially robust, these clusters are displayed up to a stricter threshold of *Z*>3.1 (*p*<0.001).

To explore associations between cerebello-cerebral connectivity and behavioral outcomes at 18 months, parameter estimates reflecting connectivity strength between the seed and the target cluster were extracted from clusters that survived correction at a threshold of *Z*>2.3. These parameter estimates were entered as predictors in linear regression models of behavioral outcomes, controlling for sex, GA at birth, PNA at scan, PNA at the time of behavioral assessment, and the first five principal ancestry components.

## Results

### Demographic and Behavioral Sex Differences

The average GA at birth was 40.18±1.21 weeks, and infants were scanned at a mean of 9.73±9.07 days postpartum. Male and female infants did not significantly differ on age, nor on ASD PGS score (Table 1). At 18 months, compared with males, females scored significantly higher on the Receptive Language subscale (BSITD-III; t=2.18, p=0.03). No other sex differences were observed on other 18-month BSITD-III measures, the Q-CHAT, or the CBCL.

### Group Connectivity Patterns

Significant cerebello-cerebral connectivity was observed for the right SensoriMotor and left SocialCognitive cerebellar seeds. The right SensoriMotor seed was functionally connected with the right inferior frontal gyrus (pars triangularis and opercularis) as well as middle temporal gyrus and temporal poles (Figure 1A). The left SocialCognitive seed had significant connectivity with a left prefrontal cluster localized to the superior frontal gyrus (dorsal and medial aspects), middle frontal gyrus, and supplementary motor area (Figure 1B). The left SensoriMotor and right SocialCognitive seeds did not exhibit connectivity exceeding the minimum significance threshold.

### Sex Differences in Connectivity

We found widespread, robust sex differences in cerebello-cerebral connectivity (*Z*>3.1) across all ROIs tested. In general, females had greater cerebellar connectivity with the medial occipital and superior temporal cortices, compared to males (Figure 2A-D). For the SensoriMotor seeds (Figure 2A-B), females showed stronger connectivity with medial prefrontal and parietal areas, while males had greater connectivity with pre– and postcentral gyri. These patterns were generally reversed for the SocialCognitive seeds (Figure 2C-D), such that females had greater synchrony between SocialCognitive seeds and sensorimotor cortices, while males had greater connectivity with medial prefrontal and parietal areas.

### Autism PGS Predicts Cerebellar Connectivity

ASD PGS predicted multiple atypical connectivity patterns. Across the entire group, infants with higher PGS showed stronger connectivity between the left SensoriMotor seed and left temporal cortex (middle and inferior temporal, as well as fusiform gyri; Figure 3A). Higher ASD PGS was also associated with overconnectivity between the right SensoriMotor seed and right parietal cortex (angular, superior parietal, and inferior parietal cortices), and weaker connectivity with bilateral posterior temporal cortex, angular and supramarginal gyri, and left inferior parietal lobule (Figure 3B). Elevated ASD PGS was also associated with hyperconnectivity between the right SocialCognitive seed and a cluster encompassing the bilateral precuneus, middle and posterior cingulate gyri, cuneus, and left calcarine cortex (Figure 3C). No clusters survived the minimum significance threshold for the left SocialCognitive seed.

Connectivity associated with elevated ASD PGS was also related to several behavioral outcomes with nominal significance (Figure 3D). In males only, stronger connectivity between the left SensoriMotor seed and temporal cortex predicted higher expressive language scores (β=5.34, p=0.026) and elevated attention problems (β=13.03, p=0.044) at 18-month outcome. Weaker connectivity between the right SensoriMotor seed and posterior temporal cortex also predicted worse attention problems in males only (β=-14.28, p=0.029). Finally, hyperconnectivity between the right SocialCognitive seed and the posterior cingulate cluster predicted reduced emotional reactivity in girls only (β=-8.60, p=0.030).

### Sex-Dependent Associations Between and Autism Polygenic Score and Cerebellar Connectivity

We observed multiple significant interactions between sex and ASD PGS, indicating sex-dependent genetic effects. For the right SensoriMotor seed, we observed significant Male>Female interactions across multiple brain regions (Figure 4A), whereas for the right SocialCognitive seed, interactions were observed in the opposite direction (Female>Male; Figure 4B).

More specifically, in males, ASD PGS was associated with stronger connectivity between the right SensoriMotor seed and the left cuneus, precuneus, and posterior cingulate with the directionality of this association reversed in females (Figure 4A & 4C-left; Male β=1.86, p<0.001; Female β=-0.87, p=0.05). Elevated ASD PGS was also associated with weaker connectivity between the right SensoriMotor seed and the right parietal cortex (angular and supramarginal gyri, as well as posterior temporal cortex) for females, but not for males (Figure 4A & 4C-middle; Female β=-1.51, p<0.001; Male β=0.92, p=0.06). Stronger connectivity between the right SensoriMotor seed and the parietal interaction cluster was also predictive of higher expressive language scores at outcome for males (Figure 4C-right; β=4.65, p=0.005). In females, ASD PGS predicted stronger connectivity between the right SocialCognitive seed and bilateral primary sensorimotor cortices, operculum, supramarginal gyrus, and left superior temporal gyrus (Figures 4B & 4D; Left cluster: β=0.88, p=0.008; Right cluster: β=0.70, p=0.033). Remarkably, this association was reversed in males such that elevated ASD PGS predicted weaker connectivity between the SocialCognitive cerebellum and these same regions (Figure 4D; Left cluster: β=-0.81, p=0.013; Right cluster: β=-0.86, p=0.005).

## Discussion

Despite the cerebellum’s clear role in learning and behavioral development, studies linking early cerebellar function to later behavioral and psychiatric outcomes are still scarce (23), and even rarer are studies that have examined how genetic variation shapes the developing brain in infancy. To address this gap, we present, to our knowledge, the first investigation of how common genetic variants associated with ASD influence functional connectivity of the infant cerebellum. Specifically, we examine how this often overlooked aspect of early brain organization varies as a function of biological sex and genetic liability for ASD in a well-powered cohort of 198 neonates with both neuroimaging and genetic data. A major limitation of prior fetal and infant neuroimaging research is the tendency to treat the cerebellum as a uniform structure, even though structural and functional subdivisions are well established in both adults (55,59) and infants (60). Here instead, we focus on anatomically defined cerebellar subregions associated with SensoriMotor and SocialCognitive behaviors. In addition to widespread sex differences in neonatal functional connectivity of the cerebellum across both SensoriMotor and SocialCognitive subdivisions, we identify significant associations between ASD polygenic scores (PGS) and subdivision-specific cerebello-cerebral connectivity. Across both sexes, elevated ASD PGS was associated with altered cerebellar connectivity, including atypical coupling between the SensoriMotor cerebellum and temporal cortex, and hyperconnectivity between the right SocialCognitive cerebellum and the posterior cingulate. Notably, brain-behavior associations were largely sex-specific, suggesting that cerebellar functional subdivisions may contribute to both sex-dependent neurodevelopment and potential compensatory mechanisms.

### Asymmetric cerebello-cerebral connectivity in neonates

We detected multiple signatures of intrinsic connectivity between the cerebellum and cortex. The right SensoriMotor seed showed significant functional synchrony with right inferior frontal and temporal areas – right-hemisphere homologs of classical language areas. Connectivity between this SensoriMotor seed and the inferior frontal gyrus (IFG) is particularly interesting, as the right IFG has also been implicated in motor inhibitory control (61), suggesting that this pattern may be a precursor to a cerebello-cerebral motor network. In contrast, the left SocialCognitive seed showed significant connectivity with the left superior and middle frontal gyri, regions of the prefrontal cortex involved in social cognition, executive function, and top-town control of implicit learning and attention (62). Notably, the left SensoriMotor and right SocialCognitive seeds lacked significant connectivity in this sample, suggesting that robust long-range cerebello-cortical connectivity is still actively developing soon after birth. Indeed, a recent study (39) demonstrated that, although intra-cerebellar connectivity may be relatively stable at birth, cerebello-cortical projections evolve dramatically within the first two years of life, indicating that long-range cerebellar connectivity is expected to be immature at this young age.

### Genetic liability for ASD alters early cerebellar-cortical network organization

Consistent with evidence implicating the cerebellum in ASD across molecular, structural, and systems levels (25), we find that genetic liability for autism shapes cerebellar functional connectivity patterns at birth. Using ASD PGS as an index of inherited genetic liability, we find that elevated ASD PGS predicts alterations in connectivity between the SensoriMotor cerebellum and supramodal cortical areas, including ipsilateral hyperconnectivity with temporal and parietal cortices. Importantly, in the mature brain, cerebello-cerebral projections decussate to contralateral cortical regions (63). Thus, our observation that infants with greater genetic liability for ASD exhibit ipsilateral overconnectivity may signal a deviation from anatomically-appropriate synchronization between the cerebellum and the cortex. Such extraneous functional synchrony may provide a mechanism early in development by which the cerebellum’s regulatory role over the cortex is perturbed in ASD. Indeed, the functional cerebellar alterations observed herein align well with the theory that ASD can be characterized as a “disorder of prediction” (30). Under this framework, the cerebellum’s predictive ability is perturbed at the cellular and transcriptomic levels (25), potentially disrupting the brain’s ability to generate reliable predictive models of environmental inputs. Compromised predictive ability may provide a unified framework for understanding the full profile of symptoms commonly observed across the autism spectrum, including difficulties in communication/language, restricted behaviors, insistence on sameness, deficits in motor coordination, and altered sensory processing.

The right SensoriMotor cerebellum additionally showed hypoconnectivity with the bilateral temporoparietal junction (TPJ) in infants with elevated PGS. This observation is intriguing given that the right cerebellum is typically implicated in language learning and processing, and that the TPJ is a multifunctional hub associated with multisensory integration, attention and cognitive control (as a key node in the ventral attention network), and higher-level language functions (64). The concurrent underengagement of these two areas in neonates may signal a neural substrate for ASD-related language and attentional difficulties, potentially reflecting issues applying cerebellar sensory feedback to shape emerging activity in the TPJ. In line with this hypothesis, we find that cerebellar-TPJ hypoconnectivity predicts a greater preponderance of attention problems in boys at 18-month outcome.

Beyond sensorimotor alterations in cerebellar functional connectivity, we also find robust hyperconnectivity between the right SocialCognitive cerebellum and a cortical cluster spanning the precuneus and posterior cingulate – key nodes of the default mode network (DMN) – for infants with elevated ASD PGS. This contrasts with previous reports of DMN-cerebellar underconnectivity in adults with (65,66). These discrepant findings may be best contextualized within their respective developmental stages, as accumulating evidence suggests that some functional connectivity differences in ASD may be life-stage dependent. Longitudinal and cross-sectional studies demonstrate that enlarged gray matter volumes in ASD detected in early childhood normalize to control levels in late childhood (67), and, similarly, within-network functional hyperconnectivity found in children with ASD resolves in adulthood (68). To date, very few studies have examined cerebellar functional connectivity in relation to ASD risk during infancy. We previously found that language-delayed infants at high likelihood for ASD show hypoconnectivity between the right crus I (part of our SocialCognitive seed) and the prefrontal cortex at 9 months of (41). We did not detect a similar cerebello-prefrontal hypoconnectivity effect in the present cohort, which may indicate that either this pattern is related more specifically to language difficulties than to broader genetic liability for ASD, or that the neonatal stage is too early in development to detect such patterns. To our knowledge, no studies have charted how cerebellar functional connectivity patterns associated with ASD unfold from infancy through early to mid childhood. Thus, it is unknown whether ASD-associated atypicalities reported in the present and prior infant cohorts persist across subsequent life stages.

### Sex-specific effects of PGS-related cerebellar connectivity on behavioral milestones

We find widespread sex differences in neonatal cerebellar functional connectivity, indicating that sexual dimorphism in cerebellar function is already present at birth. Robust sex differences in brain connectivity have been reported previously in fetal and infant studies (69,70), but, to our knowledge, no studies have focused on sex differences in *cerebellar* connectivity or examined these effects at the level of functional subdivisions. Across all cerebellar regions that we examined, females showed stronger cerebellar connectivity with medial occipital and superior temporal areas. For the SensoriMotor cerebellum, males showed stronger connectivity with pre-and postcentral gyri relative to females, while females had stronger connectivity with medial prefrontal and parietal areas. These patterns were generally reversed for the SocialCognitive cerebellum seeds. In a third-trimester fetal (mean GA: 33.6 weeks, N=118) MRI study, females showed stronger functional connectivity between the cerebellum and the left superior frontal gyrus compared to males (71), consistent with our SensoriMotor seed results. Later in development, an adolescent sample enriched for individuals with ASD showed that males have greater connectivity between the cerebellum and the default mode network relative to females (72), suggesting that sex-differences in cerebellar functional connectivity may shift across development. Sex differences in cerebellar connectivity remain understudied, and additional longitudinal work will be required to further understand how sex differences in the connectivity of cerebellar subregions may mature across the lifespan.

In our neonatal sample, elevated ASD PGS showed sex-dependent effects on cerebellar connectivity such that higher PGS was associated with opposing patterns in males and females, particularly within right-lateralized cerebellar subdivisions. For the right SensoriMotor seed, increased genetic liability for ASD was associated with stronger connectivity with posterior medial and right lateral nodes of the DMN in males, but weaker connectivity with these same regions in females. Conversely, for the right SocialCognitive seed, elevated PGS was associated with stronger connectivity with bilateral sensorimotor cortex and operculum in females and reduced connectivity in males. Sex-by-ASD diagnosis interactions have been documented for cerebellar functional connectivity in one adult study, which found that the cerebellum is globally hyperconnected to the cortex in ASD females, but hypoconnected in ASD males (73). Here, instead, we report sex differences that depend upon the cerebellar subdivision as well as the cerebral region under consideration. The cerebellum interfaces with the cortex through robust cerebello-thalamo-cortical pathways, underscoring the importance of early cerebellar feedback in shaping sensorimotor as well as associative circuits (74). Our findings suggest that this process may undergo a sexually dimorphic divergence in individuals with elevated genetic liability for ASD. The involvement of sensorimotor cortical regions is especially intriguing here, given that sensory symptoms are extremely common in ASD (75) and may arise, at least partially, from atypical connectivity between the cerebellum and the sensorimotor cortex (31). Together, these results raise the possibility that sensory circuit development may diverge by sex in infants with elevated genetic liability for ASD, potentially giving rise to sex-specific neural substrates of sensory symptoms. Longitudinal work will be important to determine how these early differences in cerebello-cerebral connectivity relate to later symptom expression.

### Limitations and Future Directions

This study has several important limitations. First, although ASD PGS provides an informative marker of common common genetic liability for autism, and thus may be especially useful for understanding the entire spectrum of autism-associated traits in the general population, we were unable to investigate associations with clinical ASD diagnoses due to the lack these data in the dHCP sample. Second, preterm-born infants were excluded in order to maintain a focused scope. However, given that the cerebellum is especially sensitive to perturbations during the perinatal period, leading to smaller cerebellar volumes (76) and altered cerebellar functional connectivity following preterm birth (38,39), future work should investigate how the effects of genetic predisposition for ASD on brain development may interact with the effects of preterm birth. Indeed, a recent review of neuroimaging work on the infant cerebellum identified no studies that concurrently examined the effects of prematurity and ASD likelihood on cerebellar features (40). Third, during the quality control stage of this project we observed a high incidence of very poor registration fit for the cerebellum. This is a common issue in infant neuroimaging pipelines and likely stems from the reality that most analytical tools are built for adult MRI data and are optimized for goodness of fit in the cerebrum rather than the cerebellum (77). Although this issue would not be a concern for most studies – which exclude the cerebellum – in the present study, this led to the necessary exclusion of nearly 25% of infants who passed all other quality control steps. Therefore, our sample size of 198 infants, while excellent amongst historical infant neuroimaging studies, is reduced relative to most other studies using dHCP data. Additionally, our sample was restricted to individuals of European ancestry because the GWAS used to generate ASD PGS included only individuals of European descent (7). While this limits the generalizability of the present results, we are hopeful that ongoing efforts to conduct multi-ancestry ASD GWAS will improve the accuracy, transferability, and equity of polygenic risk prediction across diverse populations. Finally, because the original dHCP project was cross-sectional in nature, we were unable to probe age-dependent changes in functional connectivity and behavior.

## Conclusion

To our knowledge, this is the first study to examine how common genetic variation associated with ASD influences functional connectivity of the infant cerebellum. We demonstrate widespread sex differences in neonatal cerebellar connectivity patterns, including both cerebellum-wide and subdivision-specific patterns. Elevated ASD polygenic scores were associated with altered cerebello-cerebral connectivity days after birth, with males and females showing distinct and opposing patterns of cerebellar connectivity with several cortical brain regions. These findings indicate that genetic liability for ASD does not exert uniform effects on functional brain organization but instead is instantiated in a sexually dimorphic developmental manner. Brain-behavioral associations were likewise largely sex-specific, suggesting that early compensatory or adaptive processes may depend on biological sex. Altogether, these findings indicate that polygenic liability for ASD exerts effects on the cerebellum at birth in a population-based sample, and that these early-life alterations in connectivity may have cascading impacts on later behavioral milestones and clinical symptom profiles. By integrating neuroimaging and genetics in infancy, this work advances our understanding of the early biological origins of ASD and identifies the neonatal cerebellum as a potential locus for early divergence in ASD-associated neurodevelopmental pathways.

## Data Availability

All data produced in the present study are available upon reasonable request to the authors.

## Acknowledgements

We sincerely thank all of the researchers and families who dedicated countless hours to painstakingly acquire the Developing Human Connectome Project dataset, making it publicly available to the scientific community. This work was supported by the National Library of Medicine (NLM) at the National Institutes of Health, grant number 5T15 LM013976-03 to L.W.; a UCLA Friends of Semel Scholar Grant to L.W.; the National Institute of Mental Health (NIMH) grant number 1F31 MH135704 to E.C.; NIH National Center for Advancing Translational Science (NCATS) UCLA CTSI Grant Number KL2TR001882 to J.L.; K01 MH135289 to L.M.H.; and the Simons Foundation Autism Research Initiative grant number 00011789 to L.M.H. This work was also supported by the National Institute of Child Health Development (NICHD) under Award Number P50HD103557. The content is solely the responsibility of the authors and does not necessarily represent the official views of the National Institutes of Health. The authors declare no conflicts of interest.

---

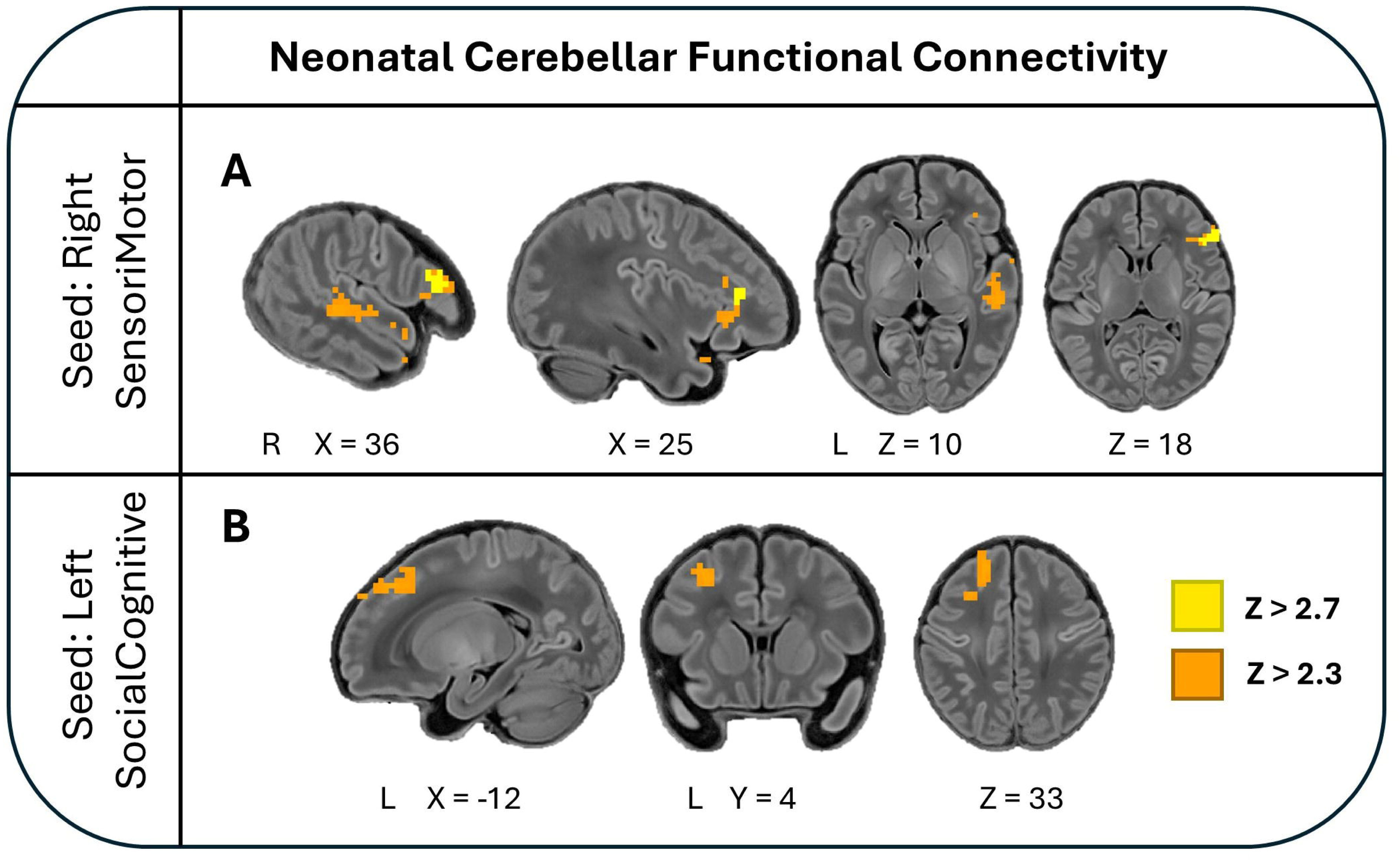

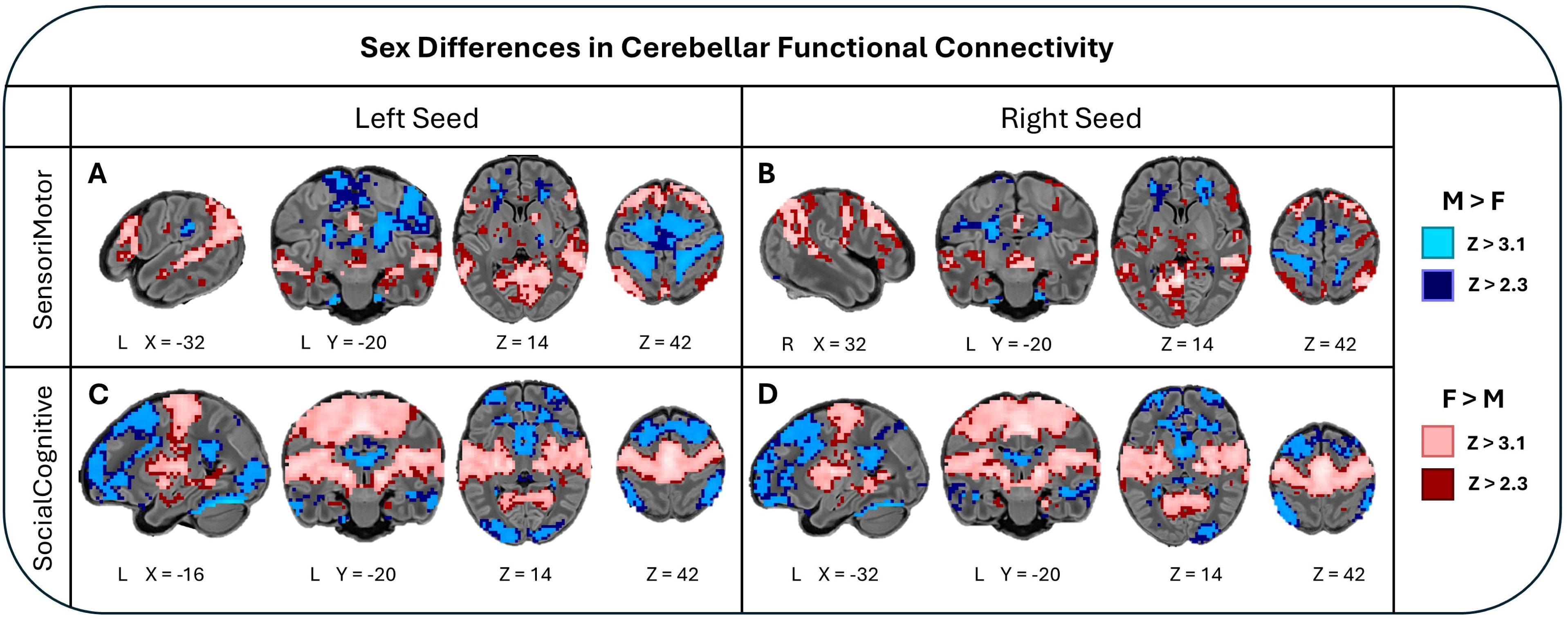

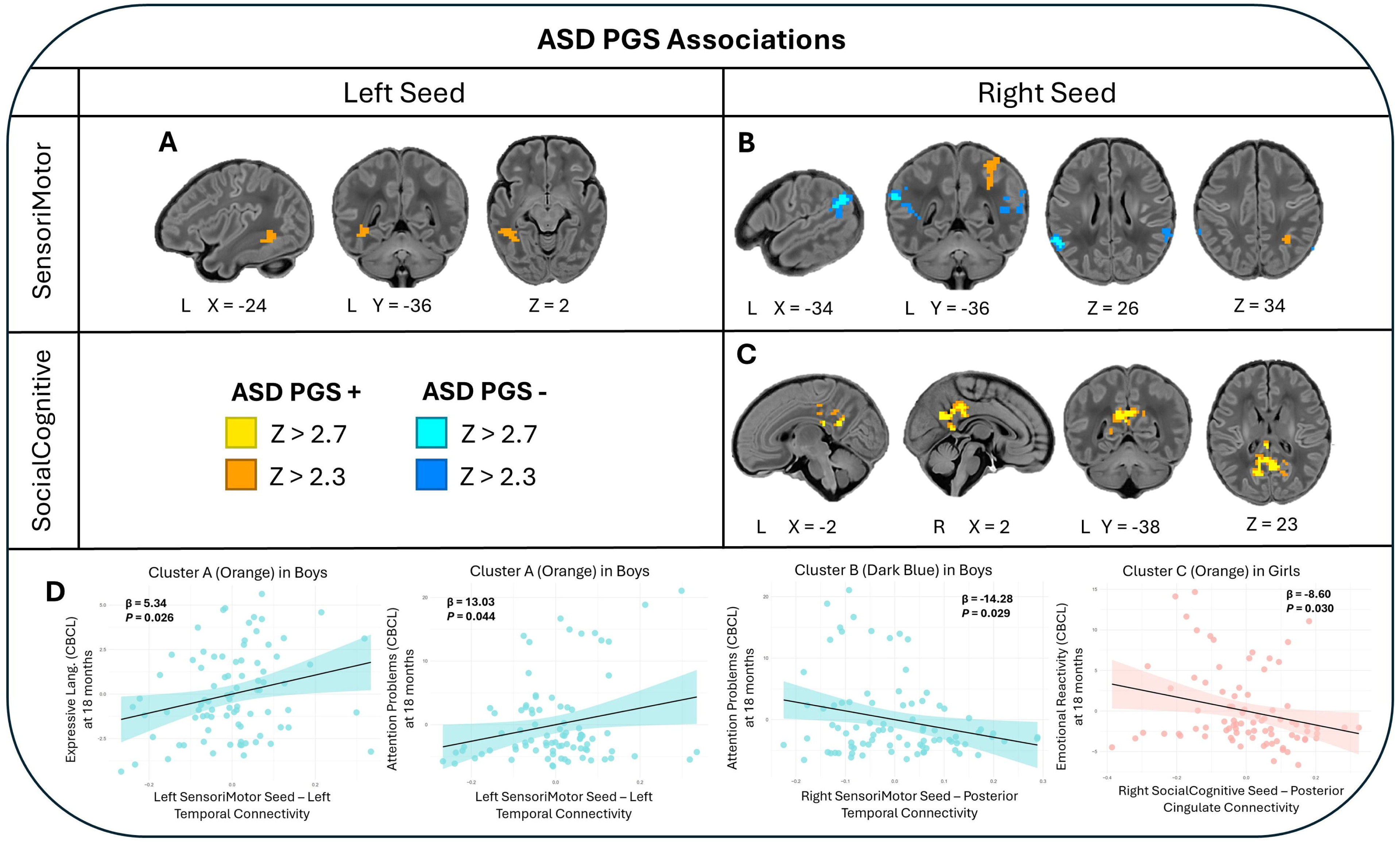

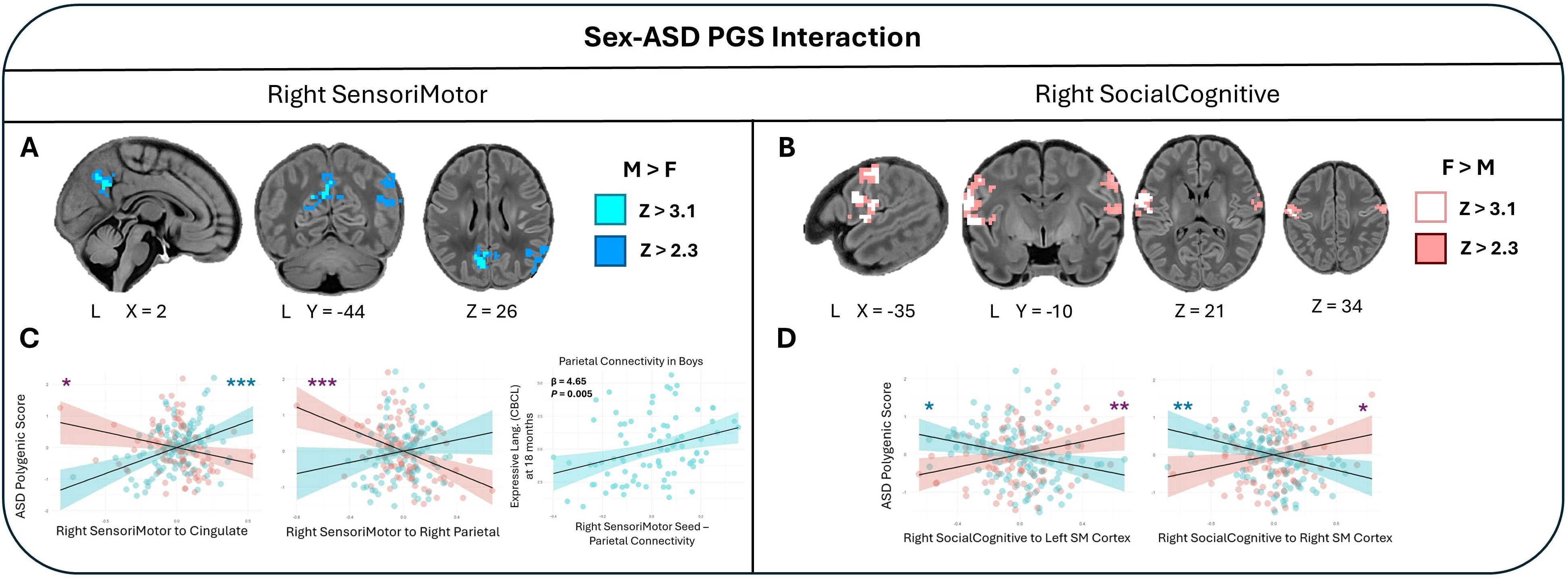

## Notes

### Competing Interest Statement

The authors have declared no competing interest.

### Author Declarations

This study used openly available human data which are publicly available for download through the NIH National Institute of Mental Health National Data Archive (https://nda.nih.gov/).

## References

1. Tick B, Bolton P, Happé F, Rutter M, Rijsdijk F. Heritability of autism spectrum disorders: a meta-analysis of twin studies. J Child Psychol Psychiatry. 2016;57(5):585–95. doi:10.1111/jcpp.12499

2. Shaw KA. Prevalence and Early Identification of Autism Spectrum Disorder Among Children Aged 4 and 8 Years — Autism and Developmental Disabilities Monitoring Network, 16 Sites, United States, 2022. MMWR Surveill Summ. 2025;74. doi:10.15585/mmwr.ss7402a1

3. Aydin E, Tsompanidis A, Chaplin D, Hawkes R, Allison C, Hackett G, et al. Fetal brain growth and infant autistic traits. Mol Autism. 2024 Feb 28;15(1):11. doi:10.1186/s13229-024-00586-5

4. Bai D, Yip BHK, Windham GC, Sourander A, Francis R, Yoffe R, et al. Association of Genetic and Environmental Factors With Autism in a 5-Country Cohort. JAMA Psychiatry. 2019 Oct;76(10):1035–43. doi:10.1001/jamapsychiatry.2019.1411 PubMed PMID: 31314057; PubMed Central PMCID: PMC6646998.

5. Anney R, Klei L, Pinto D, Almeida J, Bacchelli E, Baird G, et al. Individual common variants exert weak effects on the risk for autism spectrum disorders. Hum Mol Genet. 2012 Nov 1;21(21):4781–92. doi:10.1093/hmg/dds301

6. Gaugler T, Klei L, Sanders SJ, Bodea CA, Goldberg AP, Lee AB, et al. Most genetic risk for autism resides with common variation. Nat Genet. 2014 Aug;46(8):881–5. doi:10.1038/ng.3039

7. Grove J, Ripke S, Als TD, Mattheisen M, Walters RK, Won H, et al. Identification of common genetic risk variants for autism spectrum disorder. Nat Genet. 2019 Mar;51(3):431–44. doi:10.1038/s41588-019-0344-8

8. Klei L, Sanders SJ, Murtha MT, Hus V, Lowe JK, Willsey AJ, et al. Common genetic variants, acting additively, are a major source of risk for autism. Mol Autism. 2012 Oct 15;3(1):9. doi:10.1186/2040-2392-3-9

9. Ben-David E, Shifman S. Networks of Neuronal Genes Affected by Common and Rare Variants in Autism Spectrum Disorders. PLOS Genet. 2012 Mar 8;8(3):e1002556. doi:10.1371/journal.pgen.1002556

10. Mahfouz A, Ziats MN, Rennert OM, Lelieveldt BPF, Reinders MJT. Shared Pathways Among Autism Candidate Genes Determined by Co-expression Network Analysis of the Developing Human Brain Transcriptome. J Mol Neurosci. 2015 Dec 1;57(4):580–94. doi:10.1007/s12031-015-0641-3

11. van ‘t Hof M, Tisseur C, van Berckelear-Onnes I, van Nieuwenhuyzen A, Daniels AM, Deen M, et al. Age at autism spectrum disorder diagnosis: A systematic review and meta-analysis from 2012 to 2019. Autism. 2021 May 1;25(4):862–73. doi:10.1177/1362361320971107

12. Hyman SL, Levy SE, Myers SM, COUNCIL ON CHILDREN WITH DISABILITIES SODABP, Kuo DZ, Apkon S, et al. Identification, Evaluation, and Management of Children With Autism Spectrum Disorder. Pediatrics. 2020 Jan 1;145(1):e20193447. doi:10.1542/peds.2019-3447

13. Mohammad S, Gentreau M, Dubol M, Rukh G, Mwinyi J, Schiöth HB. Association of polygenic scores for autism with volumetric MRI phenotypes in cerebellum and brainstem in adults. Mol Autism. 2024 Aug 7;15(1):34. doi:10.1186/s13229-024-00611-7 PubMed PMID: 39113134; PubMed Central PMCID: PMC11304666.

14. Lawrence KE, Hernandez LM, Fuster E, Padgaonkar NT, Patterson G, Jung J, et al. Impact of autism genetic risk on brain connectivity: a mechanism for the female protective effect. Brain. 2022 Jan 1;145(1):378–87. doi:10.1093/brain/awab204

15. Khundrakpam B, Vainik U, Gong J, Al-Sharif N, Bhutani N, Kiar G, et al. Neural correlates of polygenic risk score for autism spectrum disorders in general population. Brain Commun. 2020 Jul 1;2(2):fcaa092. doi:10.1093/braincomms/fcaa092

16. Gu Y, Maria-Stauffer E, Bedford SA, APEX consortium, iPSYCH-autism consortium, Romero-Garcia R, et al. Polygenic scores for autism are associated with neurite density in adults and children from the general population. MedRxiv Prepr Serv Health Sci. 2024 Apr 13;2024.04.10.24305539. doi:10.1101/2024.04.10.24305539 PubMed PMID: 38645251; PubMed Central PMCID: PMC11030520.

17. Chiem E, Ganesh SSA, Dodson J, Dapretto M, Hernandez L. Effects of polygenic liability for autism on neonatal thalamocortical connectivity and behavioral outcomes across sex [Internet]. medRxiv; 2025 [cited 2026 Feb 26]. p. 2025.12.23.25342934. Available from: https://www.medrxiv.org/content/10.64898/2025.12.23.25342934v1 doi:10.64898/2025.12.23.25342934

18. Herculano-Houzel S. The human brain in numbers: a linearly scaled-up primate brain. Front Hum Neurosci. 2009 Nov 9;3. doi:10.3389/neuro.09.031.2009

19. Herculano-Houzel S. Coordinated Scaling of Cortical and Cerebellar Numbers of Neurons. Front Neuroanat. 2010 Mar 10;4:12. doi:10.3389/fnana.2010.00012 PubMed PMID: 20300467; PubMed Central PMCID: PMC2839851.

20. Baumann O, Borra RJ, Bower JM, Cullen KE, Habas C, Ivry RB, et al. Consensus Paper: The Role of the Cerebellum in Perceptual Processes. The Cerebellum. 2015 Apr 1;14(2):197–220. doi:10.1007/s12311-014-0627-7

21. LeBel A, D’Mello AM. A seat at the (language) table: incorporating the cerebellum into frameworks for language processing. Curr Opin Behav Sci. 2023 Oct 1;53:101310. doi:10.1016/j.cobeha.2023.101310

22. Schmahmann JD. The cerebellum and cognition. Neurosci Lett. 2019 Jan 1; The Cerebellum in Health and Disease 688:62–75. doi:10.1016/j.neulet.2018.07.005

23. Wagner L, Cakar ME, Banchik M, Chiem E, Glynn SS, Than AH, et al. Beyond motor learning: Insights from infant magnetic resonance imaging on the critical role of the cerebellum in behavioral development. Dev Cogn Neurosci. 2025 Apr 1;72:101514. doi:10.1016/j.dcn.2025.101514

24. D’Mello AM, Stoodley CJ. Cerebro-cerebellar circuits in autism spectrum disorder. Front Neurosci. 2015 Nov 5;9. doi:10.3389/fnins.2015.00408

25. Fatemi SH, Aldinger KA, Ashwood P, Bauman ML, Blaha CD, Blatt GJ, et al. Consensus paper: pathological role of the cerebellum in autism. Cerebellum Lond Engl. 2012 Sep;11(3):777–807. doi:10.1007/s12311-012-0355-9 PubMed PMID: 22370873; PubMed Central PMCID: PMC3677555.

26. Allen G, Müller RA, Courchesne E. Cerebellar function in autism: Functional magnetic resonance image activation during a simple motor task. Biol Psychiatry. 2004 Aug 15;56(4):269–78. doi:10.1016/j.biopsych.2004.06.005

27. Mostofsky SH, Powell SK, Simmonds DJ, Goldberg MC, Caffo B, Pekar JJ. Decreased connectivity and cerebellar activity in autism during motor task performance. Brain J Neurol. 2009 Sep;132(Pt 9):2413–25. doi:10.1093/brain/awp088 PubMed PMID: 19389870; PubMed Central PMCID: PMC2732264.

28. D’Mello AM, Moore DM, Crocetti D, Mostofsky SH, Stoodley CJ. Cerebellar gray matter differentiates children with early language delay in autism. Autism Res. 2016;9(11):1191–204. doi:10.1002/aur.1622

29. Skefos J, Cummings C, Enzer K, Holiday J, Weed K, Levy E, et al. Regional alterations in purkinje cell density in patients with autism. PloS One. 2014;9(2):e81255. doi:10.1371/journal.pone.0081255 PubMed PMID: 24586223; PubMed Central PMCID: PMC3933333.

30. Sinha P, Kjelgaard MM, Gandhi TK, Tsourides K, Cardinaux AL, Pantazis D, et al. Autism as a disorder of prediction. Proc Natl Acad Sci. 2014 Oct 21;111(42):15220–5. doi:10.1073/pnas.1416797111

31. Cakar ME, Okada NJ, Cummings KK, Jung J, Bookheimer SY, Dapretto M, et al. Functional connectivity of the sensorimotor cerebellum in autism: associations with sensory over-responsivity. Front Psychiatry. 2024 Mar 25;15. doi:10.3389/fpsyt.2024.1337921

32. Khan AJ, Nair A, Keown CL, Datko MC, Lincoln AJ, Müller RA. Cerebro-cerebellar resting state functional connectivity in children and adolescents with autism spectrum disorder. Biol Psychiatry. 2015 Nov 1;78(9):625–34. doi:10.1016/j.biopsych.2015.03.024 PubMed PMID: 25959247; PubMed Central PMCID: PMC5708535.

33. Lidstone DE, Rochowiak R, Mostofsky SH, Nebel MB. A Data Driven Approach Reveals That Anomalous Motor System Connectivity is Associated With the Severity of Core Autism Symptoms. Autism Res Off J Int Soc Autism Res. 2021 Jan 22. doi:10.1002/aur.2476 PubMed PMID: 33484109; PubMed Central PMCID: PMC8931705.

34. Arnold Anteraper S, Guell X, D’Mello A, Joshi N, Whitfield-Gabrieli S, Joshi G. Disrupted Cerebrocerebellar Intrinsic Functional Connectivity in Young Adults with High-Functioning Autism Spectrum Disorder: A Data-Driven, Whole-Brain, High-Temporal Resolution Functional Magnetic Resonance Imaging Study. Brain Connect. 2019 Feb;9(1):48–59. doi:10.1089/brain.2018.0581 PubMed PMID: 29896995.

35. Verly M, Verhoeven J, Zink I, Mantini D, Peeters R, Deprez S, et al. Altered functional connectivity of the language network in ASD: role of classical language areas and cerebellum. NeuroImage Clin. 2014;4:374–82. doi:10.1016/j.nicl.2014.01.008 PubMed PMID: 24567909; PubMed Central PMCID: PMC3930113.

36. Kelly E, Meng F, Fujita H, Morgado F, Kazemi Y, Rice LC, et al. Regulation of autism-relevant behaviors by cerebellar–prefrontal cortical circuits. Nat Neurosci. 2020 Sep;23(9):1102–10. doi:10.1038/s41593-020-0665-z

37. Kipping JA, Tuan TA, Fortier MV, Qiu A. Asynchronous Development of Cerebellar, Cerebello-Cortical, and Cortico-Cortical Functional Networks in Infancy, Childhood, and Adulthood. Cereb Cortex. 2016 Oct 12;cercor;bhw298v1. doi:10.1093/cercor/bhw298

38. Herzmann CS, Snyder AZ, Kenley JK, Rogers CE, Shimony JS, Smyser CD. Cerebellar Functional Connectivity in Term– and Very Preterm-Born Infants. Cereb Cortex. 2019 Mar 1;29(3):1174–84. doi:10.1093/cercor/bhy023

39. Tikoo S, Hernandez-Castillo CR, Chen H, Stephens R, Cornea E, Gilmore JH, et al. The Evolving Cerebellar and Cerebello-cortical Functional Connectivity Architecture during Infancy. J Neurosci. 2025 Mar 12;45(11):e1209242025. doi:10.1523/JNEUROSCI.1209-24.2025

40. Wagner L, Banchik M, Tsang T, Okada NJ, Altshuler R, McDonald N, et al. Atypical early neural responses to native and non-native language in infants at high likelihood for developing autism. Mol Autism. 2025 Feb 3;16(1):6. doi:10.1186/s13229-025-00640-w

41. Okada NJ, Liu J, Tsang T, Nosco E, McDonald NM, Cummings KK, et al. Atypical cerebellar functional connectivity at 9 months of age predicts delayed socio-communicative profiles in infants at high and low risk for autism. J Child Psychol Psychiatry. 2022;63(9):1002–16. doi:10.1111/jcpp.13555

42. Rogers SJ. What are infant siblings teaching us about autism in infancy? Autism Res Off J Int Soc Autism Res. 2009 Jun;2(3):125–37. doi:10.1002/aur.81 PubMed PMID: 19582867; PubMed Central PMCID: PMC2791538.

43. Szatmari P, Chawarska K, Dawson G, Georgiades S, Landa R, Lord C, et al. Prospective Longitudinal Studies of Infant Siblings of Children With Autism: Lessons Learned and Future Directions. J Am Acad Child Adolesc Psychiatry. 2016 Mar;55(3):179–87. doi:10.1016/j.jaac.2015.12.014 PubMed PMID: 26903251; PubMed Central PMCID: PMC4871151.

44. Ozonoff S, Young GS, Carter A, Messinger D, Yirmiya N, Zwaigenbaum L, et al. Recurrence Risk for Autism Spectrum Disorders: A Baby Siblings Research Consortium Study. Pediatrics. 2011 Sep;128(3):e488–95. doi:10.1542/peds.2010-2825 PubMed PMID: 21844053; PubMed Central PMCID: PMC3164092.

45. Ozonoff S, Young GS, Bradshaw J, Charman T, Chawarska K, Iverson JM, et al. Familial Recurrence of Autism: Updates From the Baby Siblings Research Consortium. Pediatrics. 2024 Aug 1;154(2):e2023065297. doi:10.1542/peds.2023-065297 PubMed PMID: 39011552; PubMed Central PMCID: PMC11291960.

46. Ozonoff S, Young GS, Belding A, Hill M, Hill A, Hutman T, et al. The Broader Autism Phenotype in Infancy: When Does It Emerge? J Am Acad Child Adolesc Psychiatry. 2014 Apr;53(4):398–407.e2. doi:10.1016/j.jaac.2013.12.020 PubMed PMID: 24655649; PubMed Central PMCID: PMC3989934.

47. Edwards AD, Rueckert D, Smith SM, Abo Seada S, Alansary A, Almalbis J, et al. The Developing Human Connectome Project Neonatal Data Release. Front Neurosci. 2022;16:886772. doi:10.3389/fnins.2022.886772 PubMed PMID: 35677357; PubMed Central PMCID: PMC9169090.

48. Allison C, Baron-Cohen S, Wheelwright S, Charman T, Richler J, Pasco G, et al. The Q-CHAT (Quantitative CHecklist for Autism in Toddlers): a normally distributed quantitative measure of autistic traits at 18-24 months of age: preliminary report. J Autism Dev Disord. 2008 Sep;38(8):1414–25. doi:10.1007/s10803-007-0509-7 PubMed PMID: 18240013.

49. Albers CA, Grieve AJ. Test Review: Bayley, N. (2006). Bayley Scales of Infant and Toddler Development– Third Edition. San Antonio, TX: Harcourt Assessment. J Psychoeduc Assess. 2007 Jun 1;25(2):180–90. doi:10.1177/0734282906297199

50. Achenbach TM, Ruffle TM. The Child Behavior Checklist and Related Forms for Assessing Behavioral/Emotional Problems and Competencies. Pediatr Rev. 2000 Aug 1;21(8):265–71. doi:10.1542/pir.21-8-265

51. McVean GA, Altshuler (Co-Chair) DM, Durbin (Co-Chair) RM, Abecasis GR, Bentley DR, Chakravarti A, et al. An integrated map of genetic variation from 1,092 human genomes. Nature. 2012 Nov;491(7422):56–65. doi:10.1038/nature11632

52. Ge T, Chen CY, Ni Y, Feng YCA, Smoller JW. Polygenic prediction via Bayesian regression and continuous shrinkage priors. Nat Commun. 2019 Apr 16;10(1):1776. doi:10.1038/s41467-019-09718-5

53. Fitzgibbon SP, Harrison SJ, Jenkinson M, Baxter L, Robinson EC, Bastiani M, et al. The developing Human Connectome Project (dHCP) automated resting-state functional processing framework for newborn infants. NeuroImage. 2020 Dec 1;223:117303. doi:10.1016/j.neuroimage.2020.117303

54. Smith SM, Jenkinson M, Woolrich MW, Beckmann CF, Behrens TEJ, Johansen-Berg H, et al. Advances in functional and structural MR image analysis and implementation as FSL. NeuroImage. 2004 Jan 1; Mathematics in Brain Imaging23:S208–19. doi:10.1016/j.neuroimage.2004.07.051

55. Nettekoven C, Zhi D, Shahshahani L, Pinho AL, Saadon-Grosman N, Buckner RL, et al. A hierarchical atlas of the human cerebellum for functional precision mapping. Nat Commun. 2024 Sep 27;15(1):8376. doi:10.1038/s41467-024-52371-w

56. Stoodley CJ. The Cerebellum and Cognition: Evidence from Functional Imaging Studies. The Cerebellum. 2012 Jun 1;11(2):352–65. doi:10.1007/s12311-011-0260-7

57. Stoodley CJ, Schmahmann JD. Evidence for topographic organization in the cerebellum of motor control versus cognitive and affective processing. Cortex. 2010 Jul 1; Language, Cognition and the Cerebellum: Grappling with an Enigma 46(7):831–44. doi:10.1016/j.cortex.2009.11.008

58. O’Reilly JX, Beckmann CF, Tomassini V, Ramnani N, Johansen-Berg H. Distinct and Overlapping Functional Zones in the Cerebellum Defined by Resting State Functional Connectivity. Cereb Cortex. 2010 Apr 1;20(4):953–65. doi:10.1093/cercor/bhp157

59. King M, Hernandez-Castillo CR, Poldrack RA, Ivry RB, Diedrichsen J. Functional boundaries in the human cerebellum revealed by a multi-domain task battery. Nat Neurosci. 2019 Aug;22(8):1371–8. doi:10.1038/s41593-019-0436-x

60. Lyu W, Thung KH, Huynh KM, Wang L, Lin W, Ahmad S, et al. Functional development of the human cerebellum from birth to age five. Nat Commun. 2025 Jul 10;16(1):6350. doi:10.1038/s41467-025-61465-y

61. Schaum M, Pinzuti E, Sebastian A, Lieb K, Fries P, Mobascher A, et al. Right inferior frontal gyrus implements motor inhibitory control via beta-band oscillations in humans. eLife. 10:e61679. doi:10.7554/eLife.61679 PubMed PMID: 33755019; PubMed Central PMCID: PMC8096430.

62. Pollmann S. Anterior Prefrontal Contributions to Implicit Attention Control. Brain Sci. 2012 Jun 15;2(2):254–66. doi:10.3390/brainsci2020254 PubMed PMID: 24962775; PubMed Central PMCID: PMC4061792.

63. Wilbrand FJJ. Anatomie und Physiologie der Centralgebilde des Nervensystems. Walter De Gruyter Incorporated; 1840. 218 p.

64. Bernard F, Lemee JM, Mazerand E, Leiber LM, Menei P, Ter Minassian A. The ventral attention network: the mirror of the language network in the right brain hemisphere. J Anat. 2020;237(4):632–42. doi:10.1111/joa.13223

65. Arnold Anteraper S, Guell X, D’Mello A, Joshi N, Whitfield-Gabrieli S, Joshi G. Disrupted Cerebrocerebellar Intrinsic Functional Connectivity in Young Adults with High-Functioning Autism Spectrum Disorder: A Data-Driven, Whole-Brain, High-Temporal Resolution Functional Magnetic Resonance Imaging Study. Brain Connect. 2019 Feb;9(1):48–59. doi:10.1089/brain.2018.0581

66. Joshi G, Arnold Anteraper S, Patil KR, Semwal M, Goldin RL, Furtak SL, et al. Integration and Segregation of Default Mode Network Resting-State Functional Connectivity in Transition-Age Males with High-Functioning Autism Spectrum Disorder: A Proof-of-Concept Study. Brain Connect. 2017 Nov;7(9):558–73. doi:10.1089/brain.2016.0483

67. Prigge MBD, Lange N, Bigler ED, King JB, Dean DC, Adluru N, et al. A 16-year study of longitudinal volumetric brain development in males with autism. NeuroImage. 2021 Aug 1;236:118067. doi:10.1016/j.neuroimage.2021.118067

68. Nomi JS, Uddin LQ. Developmental changes in large-scale network connectivity in autism. NeuroImage Clin. 2015 Jan 1;7:732–41. doi:10.1016/j.nicl.2015.02.024

69. Cook KM, De Asis-Cruz J, Lopez C, Quistorff J, Kapse K, Andersen N, et al. Robust sex differences in functional brain connectivity are present in utero. Cereb Cortex. 2023 Mar 10;33(6):2441–54. doi:10.1093/cercor/bhac218

70. Fenske SJ, Liu J, Chen H, Diniz MA, Stephens RL, Cornea E, et al. Sex differences in resting state functional connectivity across the first two years of life. Dev Cogn Neurosci. 2023 Apr 1;60:101235. doi:10.1016/j.dcn.2023.101235

71. Wheelock MD, Hect JL, Hernandez-Andrade E, Hassan SS, Romero R, Eggebrecht AT, et al. Sex differences in functional connectivity during fetal brain development. Dev Cogn Neurosci. 2019 Apr 1;36:100632. doi:10.1016/j.dcn.2019.100632

72. Tavares V, Fernandes LA, Antunes M, Ferreira H, Prata D. Sex Differences in Functional Connectivity Between Resting State Brain Networks in Autism Spectrum Disorder. J Autism Dev Disord. 2022 Jul 1;52(7):3088–101. doi:10.1007/s10803-021-05191-6

73. Smith REW, Avery JA, Wallace GL, Kenworthy L, Gotts SJ, Martin A. Sex Differences in Resting-State Functional Connectivity of the Cerebellum in Autism Spectrum Disorder. Front Hum Neurosci. 2019 Apr 5;13. doi:10.3389/fnhum.2019.00104

74. Palesi F, Tournier JD, Calamante F, Muhlert N, Castellazzi G, Chard D, et al. Contralateral cerebello-thalamo-cortical pathways with prominent involvement of associative areas in humans in vivo. Brain Struct Funct. 2015 Nov 1;220(6):3369–84. doi:10.1007/s00429-014-0861-2

75. Leekam SR, Nieto C, Libby SJ, Wing L, Gould J. Describing the Sensory Abnormalities of Children and Adults with Autism. J Autism Dev Disord. 2007 May 1;37(5):894–910. doi:10.1007/s10803-006-0218-7

76. Cook KM, De Asis-Cruz J, Kim JH, Basu SK, Andescavage N, Murnick J, et al. Experience of early-life pain in premature infants is associated with atypical cerebellar development and later neurodevelopmental deficits. BMC Med. 2023 Nov 14;21:435. doi:10.1186/s12916-023-03141-w PubMed PMID: 37957651; PubMed Central PMCID: PMC10644599.

77. Korom M, Camacho MC, Filippi CA, Licandro R, Moore LA, Dufford A, et al. Dear reviewers: Responses to common reviewer critiques about infant neuroimaging studies. Dev Cogn Neurosci. 2022 Feb 1;53:101055. doi:10.1016/j.dcn.2021.101055

