## Supplemental Materials for "Functional Connectivity of the Neonatal Cerebellum is Impacted by Sex and Polygenic Liability for Autism"

Supplement

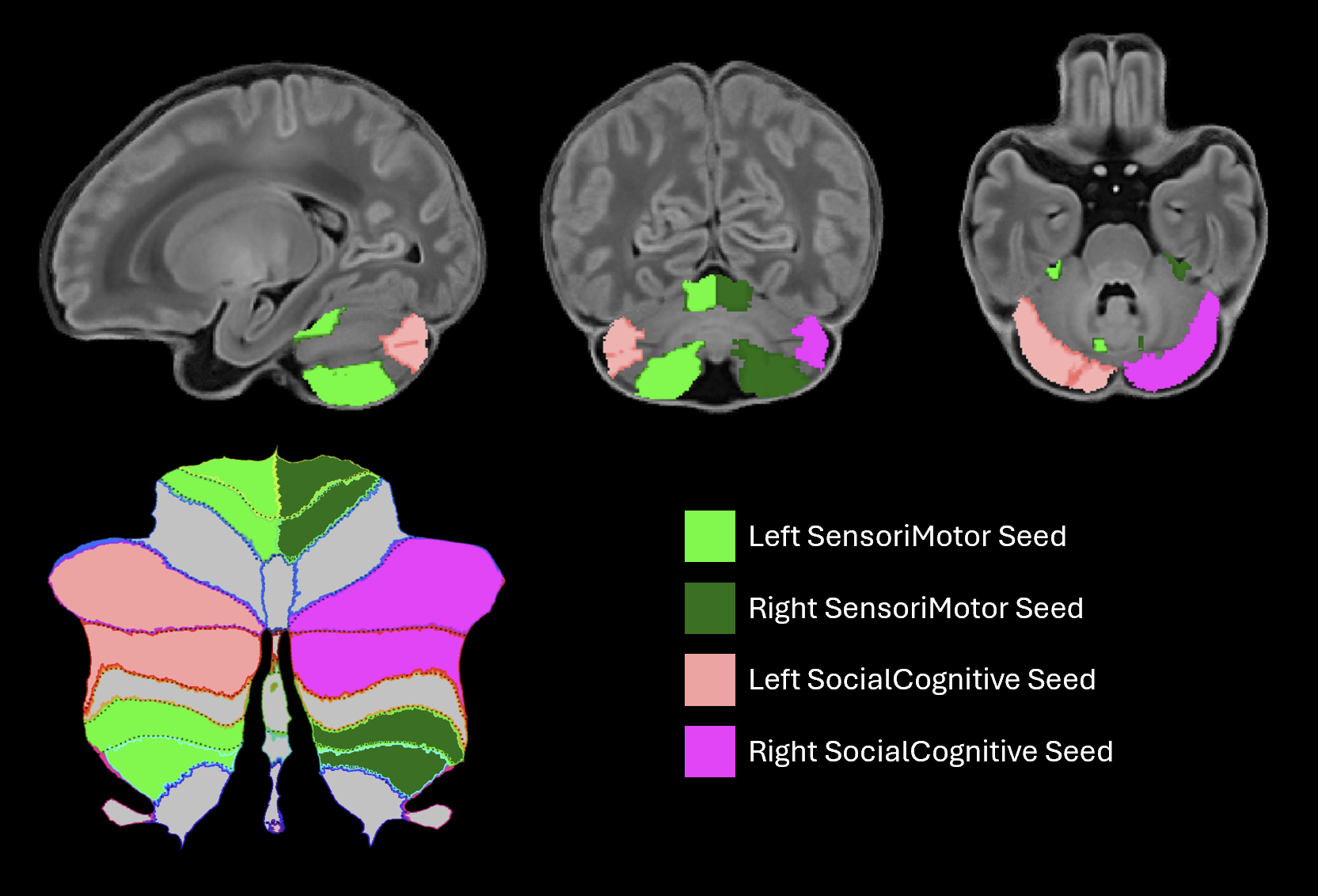

**Figure S1**. Seed regions of interest. SensoriMotor (SM) seeds are shown in green, and SocialCognitive (SC) seeds are shown in pink. A cerebellar flatmap is shown in the bottom left corner.

### Supplemental Methods

#### Participant Exclusion

Out of an original dataset of 733 infants with genetic and fMRI data, 205 were excluded due to preterm birth, 99 due to poor fMRI data quality (dHCP-rated radiology score >4 and/or failed quality control), 17 due to multiple birth to maintain independence of observations (one sibling retained in the case of twins/triplets), 141 due to non-European ancestry (incompatible with the Grove et al. (2019) GWAS), and 10 due to incomplete postnatal and/or gestational age information. During fMRI processing, an additional 63 were excluded due to poor cerebellar registration, leaving a final sample of 198 infants with quality-checked fMRI and genetic data (mean postnatal age at scan (PNA): 9.73 days, mean GA at birth: 40.18 weeks).

#### Genetic Data

Genetic data were collected through saliva samples at the initial neonatal MRI visit using Oragene DNA OG-250 kits (DNAGenotek Inc., Kanata, Canada). Biospecimens were genotyped by the NIHR Bioresource Centre Maudsley Genomics & Biomarker Core Facility using the Illumina Infinium Omni5-4 array v1.2 to identify 4,327,108 single nucleotide polymorphisms (SNPs). Genotype quality control was performed using GenomeStudio’s quality control Standard Operating Procedure (Patel et al., 2022). Samples with call rates below 95% were removed, and sex mismatches as well potential heterozygosity outliers were flagged and removed.

#### MRI Sequences

Complete information about the dHCP’s MRI sequences can be found in Edwards et al. (2022). The resting-state fMRI images were acquired using a multislice gradient-echo echo planar imaging (EPI) sequence with multiband excitation for high temporal resolution images (TR=392 ms, TE=38 ms, flip angle=34°, voxel size=2.15x2.15x2.15 mm^3^, 15 minutes 2 seconds duration, 2300 volumes). T2-weighted scans were used for anatomical co-registration (TR=12000 ms, TE=156 ms, voxel size=0.8x0.8x1.6 mm^3^, 3 minutes 12 seconds duration).

#### Supplementary References:

### Edwards AD, Rueckert D, Smith SM, Abo Seada S, Alansary A, Almalbis J, et al. The Developing Human Connectome Project Neonatal Data Release. Front Neurosci. 2022;16:886772. doi:10.3389/fnins.2022.886772 PubMed PMID: 35677357; PubMed Central PMCID: PMC9169090.

Patel H, Lee SH, Breen G, Menzel S, Ojewunmi O, Dobson RJB. The COPILOT Raw Illumina Genotyping QC Protocol. Curr Protoc. 2022;2(4):e373. doi:10.1002/cpz1.373

### Supplemental Tables

**Table S1.** MRI Coordinate Table for Group-Average Connectivity and ASD PGS associations. Voxel coordinates refer to scanner voxel coordinates. Connectivity maps were thresholded at *Z*>2.3 and cluster-corrected for multiple comparisons (*P*<0.05); only peaks that exceeded a threshold of *Z*>3.1 are reported. ASD PGS+ refers to positive associations between connectivity and ASD PGS, whereas ASD PGS- refers to negative associations.

| **Contrast** | **Seed Region** | **Structure** | **L/R** | **Max Z** | ***x*** | ***y*** | ***z*** |
| --- | --- | --- | --- | --- | --- | --- | --- |
| Group Average | SensoriMotor (Right) | Inferior frontal gyrus (opercular) | R | 4.33 | 6 | 43 | 28 |
|  |  | Inferior frontal gyrus (triangular) | R | 4.2 | 7 | 44 | 28 |
|  |  | Middle temporal gyrus | R | 3.61 | 7 | 26 | 23 |
|  |  | Orbitofrontal cortex (inferior) | R | 3.28 | 12 | 44 | 21 |
|  |  | Temporal pole (superior) | R | 3.2 | 11 | 40 | 14 |
|  |  | Insula | R | 3.18 | 12 | 43 | 21 |
|  |  | Temporal pole (middle) | R | 3.12 | 8 | 38 | 15 |
|  | SocialCognitive (Left) | Superior frontal gyrus (dorsal) | L | 4.49 | 29 | 42 | 38 |
|  |  | Supplementary motor area | L | 3.83 | 28 | 42 | 38 |
|  |  | Middle frontal gyrus | L | 3.64 | 34 | 40 | 36 |
|  |  | Superior frontal gyrus (medial) | L | 3.58 | 27 | 52 | 33 |
| ASD PGS Associations | SensoriMotor (Left) ASD PGS + | Inferior temporal gyrus | L | 3.49 | 40 | 22 | 19 |
|  |  | Middle temporal gyrus | L | 2.98 | 44 | 23 | 20 |
|  |  | Fusiform gyrus | L | 3.23 | 36 | 20 | 20 |
|  | SensoriMotor (Right) ASD PGS + | Angular gyrus | R | 3.30 | 14 | 19 | 40 |
|  |  | Superior parietal gyrus | R | 3.10 | 15 | 19 | 41 |
|  |  | Inferior parietal lobule | R | 3.33 | 11 | 24 | 41 |
|  | SensoriMotor (Right) ASD PGS - | Middle temporal gyrus | R | 3.61 | 4 | 22 | 27 |
|  |  | Superior temporal gyrus | R | 3.38 | 5 | 23 | 31 |
|  |  | Superior temporal gyrus | L | 3.48 | 43 | 20 | 30 |
|  |  | Supramarginal gyrus | L | 3.81 | 43 | 20 | 31 |
|  |  | Supramarginal gyrus | R | 2.33 | 5 | 22 | 33 |
|  |  | Angular gyrus | L | 3.59 | 42 | 17 | 32 |
|  |  | Angular gyrus | R | 2.93 | 6 | 16 | 34 |
|  |  | Middle temporal gyrus | L | 3.19 | 39 | 19 | 26 |
|  |  | Inferior parietal lobule | L | 2.73 | 44 | 22 | 35 |
|  | SocialCognitive (Right) ASD PGS + | Precuneus | L | 4.42 | 24 | 17 | 31 |
|  |  | Precuneus | R | 4.45 | 23 | 17 | 31 |
|  |  | Middle cingulate gyrus | L | 3.71 | 25 | 25 | 31 |
|  |  | Middle cingulate gyrus | R | 3.81 | 24 | 24 | 34 |
|  |  | Posterior cingulate gyrus | L | 3.73 | 26 | 19 | 30 |
|  |  | Posterior cingulate gyrus | R | 2.87 | 20 | 21 | 31 |
|  |  | Calcarine cortex | L | 2.74 | 30 | 19 | 24 |
|  |  | Cuneus | L | 3.36 | 30 | 15 | 29 |
|  |  | Cuneus | R | 2.68 | 19 | 15 | 30 |
| ASD PGS Interactions with Sex | Right SensoriMotor ASD PGS - Sex (Male > Female) | Cuneus | L | 3.91 | 27 | 14 | 31 |
|  |  | Precuneus | L | 3.43 | 25 | 17 | 32 |
|  |  | Middle temporal gyrus | R | 3.35 | 11 | 18 | 30 |
|  |  | Posterior cingulate gyrus | L | 3.24 | 28 | 20 | 30 |
|  |  | Angular gyrus | R | 3.15 | 4 | 19 | 33 |
|  |  | Superior temporal gyrus | R | 3.13 | 4 | 19 | 32 |
|  | Right SocialCognitive ASD PGS - Sex (Female > Male) | Postcentral gyrus | R | 4.14 | 5 | 32 | 36 |
|  |  | Supramarginal gyrus | R | 4.01 | 4 | 32 | 36 |
|  |  | Postcentral gyrus | L | 3.69 | 45 | 31 | 32 |
|  |  | Supramarginal gyrus | L | 3.57 | 45 | 30 | 32 |
|  |  | Superior temporal gyrus | L | 3.3 | 46 | 31 | 26 |
|  |  | Rolandic operculum | R | 3.26 | 5 | 35 | 26 |
|  |  | Rolandic operculum | L | 3.23 | 45 | 34 | 25 |
|  |  | Precentral gyrus | L | 3.16 | 42 | 36 | 30 |

**Table S2.** MRI coordinate table of sex differences in cerebellar functional connectivity. Voxel coordinates refer to scanner voxel coordinates. Connectivity maps were thresholded at *Z*>2.3 and cluster-corrected for multiple comparisons (*P*<0.05); only peaks that exceeded a threshold of *Z*>3.1 are reported.

| **Contrast** | **Seed Region** | **Structure** | **L/R** | **Max Z** | ***x*** | ***y*** | ***z*** |
| --- | --- | --- | --- | --- | --- | --- | --- |
| Male > Female | Sensorimotor (L) | Fusiform gyrus | L | 12.76 | 28 | 20 | 18 |
|  |  | Lingual gyrus | L | 11.83 | 27 | 20 | 19 |
|  |  | Parahippocampal gyrus | L | 9.64 | 29 | 24 | 16 |
|  |  | Inferior parietal lobule | R | 7.11 | 17 | 21 | 41 |
|  |  | Fusiform gyrus | R | 7.11 | 20 | 18 | 18 |
|  |  | Lingual gyrus | R | 7.06 | 21 | 19 | 19 |
|  |  | Caudate | R | 7.05 | 19 | 31 | 31 |
|  |  | Superior parietal gyrus | R | 6.95 | 18 | 21 | 41 |
|  |  | Precentral gyrus | R | 6.82 | 18 | 33 | 43 |
|  |  | Superior frontal gyrus (dorsal) | R | 6.76 | 18 | 34 | 41 |
|  |  | Precuneus | R | 6.61 | 20 | 20 | 40 |
|  |  | Superior parietal gyrus | L | 6.56 | 32 | 21 | 43 |
|  |  | Postcentral gyrus | R | 6.41 | 17 | 24 | 40 |
|  |  | Precuneus | L | 6.21 | 30 | 21 | 44 |
|  |  | Superior frontal gyrus (dorsal) | L | 6.04 | 31 | 33 | 40 |
|  |  | Inferior parietal lobule | L | 5.98 | 34 | 23 | 38 |
|  |  | Supplementary motor area | L | 5.94 | 30 | 34 | 40 |
|  |  | Thalamus | R | 5.8 | 19 | 31 | 30 |
|  |  | Supplementary motor area | R | 5.55 | 22 | 33 | 43 |
|  |  | Parahippocampal gyrus | R | 5.53 | 17 | 27 | 12 |
|  |  | Precentral gyrus | L | 5.44 | 32 | 31 | 41 |
|  |  | Postcentral gyrus | L | 5.36 | 34 | 24 | 42 |
|  |  | Supramarginal gyrus | R | 5.36 | 12 | 26 | 35 |
|  |  | Putamen | R | 5.17 | 16 | 36 | 29 |
|  |  | Middle cingulate gyrus | R | 5.15 | 24 | 35 | 37 |
|  |  | Paracentral lobule | L | 4.97 | 28 | 30 | 42 |
|  |  | Insula | R | 4.78 | 15 | 35 | 29 |
|  |  | Middle cingulate gyrus | L | 4.73 | 29 | 36 | 35 |
|  |  | Caudate | L | 4.66 | 31 | 28 | 30 |
|  |  | Paracentral lobule | R | 4.65 | 19 | 24 | 39 |
|  |  | Thalamus | L | 4.64 | 28 | 29 | 27 |
|  |  | Angular gyrus | R | 4.42 | 17 | 19 | 40 |
|  |  | Middle frontal gyrus | R | 4.4 | 16 | 35 | 41 |
|  |  | Middle frontal gyrus | L | 4.39 | 33 | 34 | 40 |
|  |  | Supramarginal gyrus | L | 4.38 | 41 | 25 | 31 |
|  |  | Pallidum | R | 4.16 | 17 | 30 | 25 |
|  |  | Rolandic operculum | R | 4.04 | 12 | 26 | 32 |
|  |  | Orbitofrontal cortex (middle) | L | 4.02 | 31 | 51 | 17 |
|  |  | Orbitofrontal cortex (superior) | L | 3.91 | 30 | 51 | 17 |
|  |  | Orbitofrontal cortex (middle) | R | 3.86 | 14 | 51 | 18 |
|  |  | Rolandic operculum | L | 3.86 | 36 | 25 | 32 |
|  |  | Orbitofrontal cortex (medial) | L | 3.76 | 28 | 51 | 17 |
|  |  | Anterior cingulate gyrus | L | 3.75 | 29 | 47 | 23 |
|  |  | Superior temporal gyrus | L | 3.72 | 40 | 26 | 30 |
|  |  | Putamen | L | 3.71 | 33 | 38 | 22 |
|  |  | Superior temporal gyrus | R | 3.7 | 6 | 27 | 31 |
|  |  | Inferior frontal gyrus (triangular) | R | 3.64 | 14 | 42 | 28 |
|  |  | Rectus gyrus | L | 3.52 | 28 | 50 | 16 |
|  |  | Inferior frontal gyrus (triangular) | L | 3.47 | 34 | 48 | 24 |
|  |  | Orbitofrontal cortex (inferior) | L | 3.46 | 32 | 45 | 20 |
|  |  | Orbitofrontal cortex (superior) | R | 3.46 | 19 | 50 | 17 |
|  |  | Insula | L | 3.46 | 33 | 41 | 25 |
|  |  | Orbitofrontal cortex (inferior) | R | 3.44 | 13 | 48 | 21 |
|  |  | Superior occipital gyrus | R | 3.38 | 19 | 17 | 38 |
|  |  | Superior frontal gyrus (medial) | L | 3.35 | 29 | 51 | 23 |
|  |  | Middle occipital gyrus | L | 3.33 | 31 | 18 | 35 |
|  |  | Inferior frontal gyrus (opercular) | R | 3.17 | 13 | 41 | 28 |
|  | Sensorimotor (R) | Fusiform gyrus | R | 14.04 | 20 | 18 | 18 |
|  |  | Lingual gyrus | R | 13.09 | 21 | 18 | 19 |
|  |  | Fusiform gyrus | L | 7.23 | 28 | 20 | 18 |
|  |  | Superior parietal gyrus | L | 6.92 | 32 | 21 | 43 |
|  |  | Parahippocampal gyrus | R | 6.8 | 17 | 27 | 12 |
|  |  | Precuneus | L | 6.5 | 30 | 18 | 43 |
|  |  | Caudate | L | 5.96 | 30 | 31 | 30 |
|  |  | Lingual gyrus | L | 5.92 | 27 | 20 | 19 |
|  |  | Superior frontal gyrus (dorsal) | R | 5.53 | 18 | 34 | 42 |
|  |  | Superior frontal gyrus (dorsal) | L | 5.52 | 32 | 33 | 40 |
|  |  | Inferior parietal lobule | L | 5.5 | 37 | 22 | 42 |
|  |  | Thalamus | L | 5.45 | 30 | 30 | 30 |
|  |  | Parahippocampal gyrus | L | 5.36 | 31 | 26 | 13 |
|  |  | Postcentral gyrus | L | 5.36 | 35 | 24 | 42 |
|  |  | Middle frontal gyrus | L | 5.31 | 33 | 46 | 27 |
|  |  | Superior parietal gyrus | R | 5.3 | 20 | 18 | 44 |
|  |  | Middle cingulate gyrus | R | 5.19 | 18 | 26 | 35 |
|  |  | Supplementary motor area | R | 5.17 | 21 | 36 | 41 |
|  |  | Supplementary motor area | L | 5.05 | 30 | 34 | 40 |
|  |  | Orbitofrontal cortex (middle) | R | 4.83 | 17 | 48 | 20 |
|  |  | Precentral gyrus | L | 4.83 | 32 | 32 | 38 |
|  |  | Caudate | R | 4.81 | 18 | 34 | 32 |
|  |  | Inferior parietal lobule | R | 4.8 | 16 | 20 | 41 |
|  |  | Inferior frontal gyrus (triangular) | L | 4.8 | 34 | 48 | 24 |
|  |  | Precuneus | R | 4.65 | 19 | 19 | 37 |
|  |  | Superior temporal gyrus | L | 4.65 | 40 | 26 | 31 |
|  |  | Thalamus | R | 4.55 | 18 | 28 | 30 |
|  |  | Supramarginal gyrus | L | 4.53 | 39 | 26 | 31 |
|  |  | Middle cingulate gyrus | L | 4.47 | 29 | 35 | 36 |
|  |  | Insula | L | 4.36 | 34 | 33 | 30 |
|  |  | Insula | R | 4.28 | 16 | 42 | 26 |
|  |  | Middle frontal gyrus | R | 4.22 | 15 | 48 | 26 |
|  |  | Precentral gyrus | R | 4.22 | 18 | 35 | 38 |
|  |  | Superior occipital gyrus | R | 4.19 | 18 | 18 | 36 |
|  |  | Putamen | L | 4.14 | 33 | 35 | 29 |
|  |  | Orbitofrontal cortex (inferior) | R | 4.1 | 17 | 47 | 20 |
|  |  | Putamen | R | 4.02 | 16 | 36 | 29 |
|  |  | Rolandic operculum | L | 4.01 | 39 | 27 | 31 |
|  |  | Paracentral lobule | L | 3.91 | 28 | 31 | 44 |
|  |  | Middle occipital gyrus | L | 3.75 | 31 | 18 | 36 |
|  |  | Inferior frontal gyrus (opercular) | L | 3.68 | 35 | 36 | 30 |
|  |  | Postcentral gyrus | R | 3.65 | 17 | 23 | 39 |
|  |  | Orbitofrontal cortex (middle) | L | 3.64 | 32 | 51 | 18 |
|  |  | Anterior cingulate gyrus | L | 3.54 | 28 | 46 | 23 |
|  |  | Angular gyrus | R | 3.41 | 17 | 19 | 35 |
|  |  | Orbitofrontal cortex (superior) | R | 3.35 | 18 | 48 | 19 |
|  |  | Orbitofrontal cortex (inferior) | L | 3.31 | 34 | 49 | 17 |
|  |  | Cuneus | R | 3.24 | 18 | 19 | 34 |
|  |  | Rolandic operculum | R | 3.14 | 11 | 38 | 27 |
|  | SocialCognitive (L) | Inferior temporal gyrus | L | 8.08 | 37 | 20 | 13 |
|  |  | Thalamus | R | 8.02 | 23 | 33 | 28 |
|  |  | Thalamus | L | 7.64 | 25 | 31 | 28 |
|  |  | Middle cingulate gyrus | R | 6.95 | 24 | 45 | 33 |
|  |  | Fusiform gyrus | R | 6.76 | 14 | 18 | 15 |
|  |  | Temporal pole (superior) | R | 6.75 | 9 | 41 | 21 |
|  |  | Caudate | R | 6.69 | 18 | 38 | 31 |
|  |  | Fusiform gyrus | L | 6.62 | 35 | 20 | 14 |
|  |  | Inferior frontal gyrus (opercular) | R | 6.61 | 8 | 41 | 22 |
|  |  | Superior frontal gyrus (medial) | L | 6.44 | 25 | 45 | 36 |
|  |  | Inferior parietal lobule | R | 6.34 | 9 | 21 | 38 |
|  |  | Caudate | L | 6.22 | 31 | 37 | 30 |
|  |  | Inferior temporal gyrus | R | 6.13 | 12 | 15 | 15 |
|  |  | Middle frontal gyrus | R | 6.12 | 15 | 53 | 27 |
|  |  | Orbitofrontal cortex (inferior) | L | 5.91 | 32 | 46 | 20 |
|  |  | Orbitofrontal cortex (inferior) | R | 5.9 | 10 | 41 | 21 |
|  |  | Insula | L | 5.89 | 39 | 40 | 21 |
|  |  | Angular gyrus | L | 5.87 | 38 | 19 | 35 |
|  |  | Supplementary motor area | L | 5.84 | 25 | 44 | 37 |
|  |  | Angular gyrus | R | 5.74 | 8 | 19 | 36 |
|  |  | Middle temporal gyrus | L | 5.73 | 43 | 24 | 19 |
|  |  | Inferior frontal gyrus (triangular) | R | 5.69 | 9 | 42 | 22 |
|  |  | Middle cingulate gyrus | L | 5.64 | 25 | 43 | 33 |
|  |  | Middle frontal gyrus | L | 5.52 | 36 | 38 | 35 |
|  |  | Superior frontal gyrus (medial) | R | 5.51 | 24 | 45 | 35 |
|  |  | Insula | R | 5.47 | 10 | 40 | 21 |
|  |  | Supramarginal gyrus | R | 5.4 | 11 | 23 | 34 |
|  |  | Precentral gyrus | R | 5.38 | 14 | 37 | 39 |
|  |  | Anterior cingulate gyrus | L | 5.33 | 25 | 45 | 32 |
|  |  | Inferior parietal lobule | L | 5.28 | 38 | 21 | 35 |
|  |  | Anterior cingulate gyrus | R | 5.23 | 24 | 45 | 31 |
|  |  | Temporal pole (superior) | L | 5.19 | 40 | 40 | 21 |
|  |  | Inferior frontal gyrus (triangular) | L | 5.18 | 39 | 40 | 22 |
|  |  | Superior occipital gyrus | L | 5.18 | 29 | 4 | 25 |
|  |  | Orbitofrontal cortex (middle) | L | 5.15 | 31 | 49 | 17 |
|  |  | Superior frontal gyrus (dorsal) | R | 5.1 | 18 | 38 | 41 |
|  |  | Middle occipital gyrus | L | 5.08 | 33 | 8 | 24 |
|  |  | Supplementary motor area | R | 4.97 | 23 | 42 | 39 |
|  |  | Orbitofrontal cortex (superior) | L | 4.95 | 30 | 50 | 18 |
|  |  | Precentral gyrus | L | 4.92 | 38 | 38 | 36 |
|  |  | Lingual gyrus | L | 4.9 | 34 | 9 | 18 |
|  |  | Inferior frontal gyrus (opercular) | L | 4.7 | 40 | 40 | 22 |
|  |  | Middle occipital gyrus | R | 4.69 | 17 | 6 | 24 |
|  |  | Superior frontal gyrus (dorsal) | L | 4.68 | 31 | 37 | 38 |
|  |  | Orbitofrontal cortex (middle) | R | 4.67 | 18 | 50 | 15 |
|  |  | Hippocampus | R | 4.66 | 13 | 24 | 22 |
|  |  | Inferior occipital gyrus | L | 4.64 | 34 | 10 | 19 |
|  |  | Middle temporal gyrus | R | 4.61 | 5 | 26 | 18 |
|  |  | Calcarine cortex | L | 4.48 | 30 | 6 | 21 |
|  |  | Lingual gyrus | R | 4.42 | 19 | 12 | 17 |
|  |  | Pallidum | R | 4.36 | 21 | 37 | 23 |
|  |  | Superior occipital gyrus | R | 4.32 | 18 | 6 | 26 |
|  |  | Cuneus | R | 4.25 | 19 | 6 | 26 |
|  |  | Parahippocampal gyrus | L | 4.01 | 35 | 22 | 21 |
|  |  | Cuneus | L | 3.98 | 28 | 4 | 28 |
|  |  | Parahippocampal gyrus | R | 3.98 | 17 | 27 | 12 |
|  |  | Rolandic operculum | R | 3.91 | 9 | 39 | 23 |
|  |  | Precuneus | R | 3.8 | 18 | 20 | 34 |
|  |  | Calcarine cortex | R | 3.69 | 20 | 6 | 24 |
|  |  | Hippocampus | L | 3.59 | 36 | 26 | 19 |
|  |  | Supramarginal gyrus | L | 3.56 | 39 | 22 | 34 |
|  |  | Pallidum | L | 3.41 | 28 | 37 | 24 |
|  |  | Inferior occipital gyrus | R | 3.39 | 12 | 10 | 23 |
|  |  | Superior parietal gyrus | L | 3.35 | 36 | 15 | 40 |
|  |  | Putamen | R | 3.26 | 20 | 38 | 23 |
|  |  | Rolandic operculum | L | 3.25 | 42 | 39 | 22 |
|  |  | Posterior cingulate gyrus | L | 3.24 | 27 | 25 | 31 |
|  |  | Putamen | L | 3.24 | 30 | 37 | 23 |
|  |  | Orbitofrontal cortex (superior) | R | 3.22 | 18 | 46 | 19 |
|  | SocialCognitive (R) | Inferior temporal gyrus | R | 10.12 | 12 | 15 | 15 |
|  |  | Fusiform gyrus | R | 9.74 | 14 | 18 | 15 |
|  |  | Lingual gyrus | R | 7.48 | 16 | 10 | 16 |
|  |  | Thalamus | L | 7.35 | 25 | 32 | 28 |
|  |  | Angular gyrus | L | 7.03 | 38 | 19 | 34 |
|  |  | Caudate | L | 6.57 | 31 | 38 | 31 |
|  |  | Caudate | R | 6.19 | 18 | 41 | 29 |
|  |  | Middle frontal gyrus | R | 6.18 | 15 | 53 | 27 |
|  |  | Superior frontal gyrus (medial) | L | 6.05 | 26 | 45 | 35 |
|  |  | Inferior parietal lobule | L | 5.96 | 38 | 21 | 34 |
|  |  | Insula | L | 5.85 | 38 | 39 | 21 |
|  |  | Middle temporal gyrus | R | 5.77 | 6 | 25 | 21 |
|  |  | Temporal pole (superior) | L | 5.72 | 39 | 40 | 19 |
|  |  | Middle frontal gyrus | L | 5.7 | 35 | 42 | 34 |
|  |  | Thalamus | R | 5.63 | 23 | 33 | 28 |
|  |  | Orbitofrontal cortex (inferior) | L | 5.6 | 38 | 40 | 19 |
|  |  | Precentral gyrus | L | 5.59 | 38 | 38 | 36 |
|  |  | Hippocampus | R | 5.43 | 12 | 28 | 19 |
|  |  | Insula | R | 5.36 | 14 | 41 | 28 |
|  |  | Middle temporal gyrus | L | 5.21 | 38 | 18 | 30 |
|  |  | Supramarginal gyrus | L | 5.12 | 38 | 22 | 33 |
|  |  | Superior parietal gyrus | L | 5.09 | 36 | 15 | 40 |
|  |  | Middle cingulate gyrus | L | 4.98 | 26 | 42 | 34 |
|  |  | Inferior parietal lobule | R | 4.96 | 8 | 19 | 37 |
|  |  | Inferior frontal gyrus (opercular) | R | 4.94 | 13 | 41 | 28 |
|  |  | Superior frontal gyrus (dorsal) | L | 4.92 | 31 | 42 | 37 |
|  |  | Inferior occipital gyrus | R | 4.91 | 14 | 11 | 19 |
|  |  | Temporal pole (superior) | R | 4.91 | 9 | 41 | 21 |
|  |  | Inferior frontal gyrus (triangular) | R | 4.85 | 13 | 44 | 26 |
|  |  | Angular gyrus | R | 4.85 | 8 | 19 | 36 |
|  |  | Olfactory cortex | L | 4.82 | 26 | 41 | 23 |
|  |  | Orbitofrontal cortex (middle) | L | 4.76 | 32 | 49 | 16 |
|  |  | Orbitofrontal cortex (superior) | L | 4.7 | 30 | 53 | 18 |
|  |  | Middle occipital gyrus | R | 4.69 | 16 | 7 | 23 |
|  |  | Supplementary motor area | L | 4.67 | 26 | 42 | 36 |
|  |  | Orbitofrontal cortex (inferior) | R | 4.66 | 17 | 47 | 20 |
|  |  | Inferior frontal gyrus (opercular) | L | 4.63 | 41 | 40 | 23 |
|  |  | Parahippocampal gyrus | R | 4.53 | 13 | 23 | 21 |
|  |  | Middle cingulate gyrus | R | 4.52 | 24 | 45 | 34 |
|  |  | Calcarine cortex | R | 4.5 | 17 | 7 | 23 |
|  |  | Anterior cingulate gyrus | L | 4.42 | 26 | 47 | 29 |
|  |  | Superior frontal gyrus (medial) | R | 4.35 | 24 | 46 | 35 |
|  |  | Inferior temporal gyrus | L | 4.34 | 37 | 27 | 18 |
|  |  | Inferior frontal gyrus (triangular) | L | 4.24 | 39 | 40 | 23 |
|  |  | Pallidum | R | 4.22 | 21 | 36 | 24 |
|  |  | Calcarine cortex | L | 4.21 | 30 | 6 | 22 |
|  |  | Supramarginal gyrus | R | 4.15 | 11 | 23 | 34 |
|  |  | Superior occipital gyrus | R | 4.14 | 20 | 3 | 28 |
|  |  | Middle occipital gyrus | L | 4.12 | 36 | 13 | 35 |
|  |  | Orbitofrontal cortex (middle) | R | 4.09 | 18 | 53 | 17 |
|  |  | Superior frontal gyrus (dorsal) | R | 4.07 | 19 | 39 | 38 |
|  |  | Precentral gyrus | R | 3.83 | 16 | 37 | 38 |
|  |  | Inferior occipital gyrus | L | 3.77 | 40 | 14 | 16 |
|  |  | Superior occipital gyrus | L | 3.68 | 30 | 7 | 23 |
|  |  | Anterior cingulate gyrus | R | 3.58 | 24 | 40 | 29 |
|  |  | Fusiform gyrus | L | 3.54 | 32 | 26 | 12 |
|  |  | Supplementary motor area | R | 3.5 | 24 | 43 | 37 |
|  |  | Cuneus | R | 3.47 | 19 | 7 | 26 |
|  |  | Orbitofrontal cortex (superior) | R | 3.43 | 18 | 47 | 19 |
|  |  | Rectus gyrus | L | 3.41 | 28 | 40 | 18 |
|  |  | Hippocampus | L | 3.38 | 36 | 25 | 20 |
|  |  | Pallidum | L | 3.35 | 28 | 35 | 25 |
|  |  | Lingual gyrus | L | 3.34 | 31 | 9 | 21 |
| Female > Male | Sensorimotor (L) | Posterior cingulate gyrus | L | 7.07 | 25 | 22 | 31 |
|  |  | Middle cingulate gyrus | R | 6.81 | 24 | 23 | 31 |
|  |  | Cuneus | R | 6.64 | 22 | 13 | 33 |
|  |  | Middle temporal gyrus | R | 6.58 | 4 | 21 | 24 |
|  |  | Middle temporal gyrus | L | 6.44 | 44 | 25 | 22 |
|  |  | Precuneus | L | 6.42 | 26 | 18 | 32 |
|  |  | Cuneus | L | 6.35 | 27 | 13 | 32 |
|  |  | Precuneus | R | 6.04 | 22 | 13 | 34 |
|  |  | Middle cingulate gyrus | L | 5.93 | 25 | 30 | 34 |
|  |  | Posterior cingulate gyrus | R | 5.88 | 23 | 23 | 30 |
|  |  | Calcarine cortex | R | 5.84 | 24 | 16 | 27 |
|  |  | Lingual gyrus | R | 5.82 | 22 | 20 | 23 |
|  |  | Inferior frontal gyrus (triangular) | R | 5.59 | 9 | 44 | 30 |
|  |  | Inferior frontal gyrus (triangular) | L | 5.54 | 38 | 41 | 30 |
|  |  | Middle frontal gyrus | R | 5.35 | 14 | 43 | 37 |
|  |  | Middle frontal gyrus | L | 5.33 | 32 | 43 | 38 |
|  |  | Angular gyrus | L | 5.28 | 38 | 12 | 37 |
|  |  | Superior temporal gyrus | R | 5.26 | 5 | 34 | 20 |
|  |  | Calcarine cortex | L | 5.25 | 24 | 16 | 25 |
|  |  | Precentral gyrus | R | 5.22 | 11 | 40 | 38 |
|  |  | Precentral gyrus | L | 5.05 | 38 | 37 | 32 |
|  |  | Lingual gyrus | L | 5.03 | 24 | 16 | 24 |
|  |  | Superior frontal gyrus (medial) | L | 4.89 | 26 | 43 | 35 |
|  |  | Orbitofrontal cortex (inferior) | R | 4.82 | 10 | 43 | 20 |
|  |  | Amygdala | L | 4.82 | 33 | 32 | 20 |
|  |  | Supplementary motor area | L | 4.8 | 26 | 41 | 38 |
|  |  | Inferior frontal gyrus (opercular) | R | 4.78 | 7 | 42 | 24 |
|  |  | Inferior parietal lobule | L | 4.78 | 35 | 14 | 38 |
|  |  | Superior frontal gyrus (dorsal) | L | 4.76 | 30 | 45 | 38 |
|  |  | Temporal pole (superior) | R | 4.72 | 13 | 42 | 15 |
|  |  | Inferior frontal gyrus (opercular) | L | 4.71 | 37 | 37 | 32 |
|  |  | Hippocampus | R | 4.67 | 13 | 29 | 19 |
|  |  | Temporal pole (middle) | R | 4.66 | 7 | 38 | 16 |
|  |  | Angular gyrus | R | 4.65 | 10 | 17 | 35 |
|  |  | Superior frontal gyrus (dorsal) | R | 4.64 | 18 | 47 | 37 |
|  |  | Orbitofrontal cortex (inferior) | L | 4.54 | 37 | 42 | 21 |
|  |  | Insula | L | 4.42 | 37 | 33 | 25 |
|  |  | Postcentral gyrus | R | 4.39 | 3 | 35 | 32 |
|  |  | Superior occipital gyrus | R | 4.38 | 17 | 13 | 28 |
|  |  | Superior frontal gyrus (medial) | R | 4.37 | 24 | 44 | 35 |
|  |  | Middle occipital gyrus | L | 4.33 | 40 | 11 | 31 |
|  |  | Putamen | L | 4.31 | 34 | 31 | 20 |
|  |  | Insula | R | 4.3 | 11 | 42 | 21 |
|  |  | Inferior temporal gyrus | R | 4.27 | 8 | 35 | 10 |
|  |  | Hippocampus | L | 4.23 | 31 | 22 | 24 |
|  |  | Putamen | R | 4.22 | 19 | 37 | 24 |
|  |  | Pallidum | R | 4.19 | 20 | 37 | 24 |
|  |  | Superior parietal gyrus | L | 4.12 | 32 | 10 | 39 |
|  |  | Caudate | R | 4.1 | 20 | 38 | 24 |
|  |  | Superior temporal gyrus | L | 4.03 | 44 | 27 | 23 |
|  |  | Supplementary motor area | R | 4.03 | 23 | 44 | 38 |
|  |  | Anterior cingulate gyrus | L | 4 | 26 | 45 | 30 |
|  |  | Supramarginal gyrus | R | 3.99 | 3 | 32 | 32 |
|  |  | Superior parietal gyrus | R | 3.96 | 23 | 11 | 38 |
|  |  | Fusiform gyrus | R | 3.94 | 17 | 20 | 19 |
|  |  | Pallidum | L | 3.94 | 29 | 34 | 24 |
|  |  | Postcentral gyrus | L | 3.88 | 45 | 32 | 33 |
|  |  | Parahippocampal gyrus | R | 3.82 | 16 | 22 | 22 |
|  |  | Inferior temporal gyrus | L | 3.72 | 43 | 27 | 15 |
|  |  | Thalamus | L | 3.7 | 27 | 29 | 22 |
|  |  | Anterior cingulate gyrus | R | 3.66 | 23 | 45 | 30 |
|  |  | Caudate | L | 3.65 | 28 | 39 | 27 |
|  |  | Superior occipital gyrus | L | 3.64 | 32 | 10 | 38 |
|  |  | Inferior parietal lobule | R | 3.63 | 8 | 18 | 37 |
|  |  | Thalamus | R | 3.56 | 22 | 29 | 23 |
|  |  | Supramarginal gyrus | L | 3.48 | 45 | 32 | 35 |
|  |  | Rolandic operculum | R | 3.47 | 13 | 33 | 28 |
|  |  | Middle occipital gyrus | R | 3.27 | 9 | 14 | 32 |
|  |  | Rolandic operculum | L | 3.17 | 37 | 32 | 28 |
|  | Sensorimotor (R) | Posterior cingulate gyrus | L | 6 | 25 | 25 | 30 |
|  |  | Calcarine cortex | L | 5.97 | 25 | 17 | 25 |
|  |  | Middle cingulate gyrus | L | 5.67 | 25 | 25 | 31 |
|  |  | Precuneus | L | 5.63 | 26 | 19 | 32 |
|  |  | Cuneus | L | 5.63 | 29 | 16 | 28 |
|  |  | Middle cingulate gyrus | R | 5.5 | 24 | 24 | 30 |
|  |  | Precuneus | R | 5.12 | 22 | 19 | 32 |
|  |  | Cuneus | R | 5.09 | 22 | 12 | 35 |
|  |  | Precentral gyrus | R | 5.07 | 11 | 37 | 34 |
|  |  | Calcarine cortex | R | 5.02 | 24 | 17 | 25 |
|  |  | Angular gyrus | R | 4.86 | 10 | 16 | 39 |
|  |  | Posterior cingulate gyrus | R | 4.83 | 24 | 20 | 28 |
|  |  | Lingual gyrus | L | 4.75 | 31 | 21 | 22 |
|  |  | Inferior frontal gyrus (triangular) | R | 4.72 | 8 | 44 | 31 |
|  |  | Inferior frontal gyrus (opercular) | R | 4.67 | 10 | 40 | 33 |
|  |  | Fusiform gyrus | L | 4.67 | 32 | 12 | 19 |
|  |  | Middle frontal gyrus | R | 4.66 | 11 | 42 | 36 |
|  |  | Middle temporal gyrus | R | 4.64 | 10 | 23 | 25 |
|  |  | Orbitofrontal cortex (medial) | L | 4.62 | 25 | 47 | 17 |
|  |  | Postcentral gyrus | R | 4.56 | 3 | 35 | 32 |
|  |  | Superior frontal gyrus (dorsal) | L | 4.47 | 30 | 45 | 38 |
|  |  | Postcentral gyrus | L | 4.41 | 43 | 33 | 29 |
|  |  | Orbitofrontal cortex (medial) | R | 4.39 | 23 | 48 | 17 |
|  |  | Middle frontal gyrus | L | 4.38 | 35 | 41 | 40 |
|  |  | Hippocampus | L | 4.34 | 34 | 30 | 19 |
|  |  | Superior frontal gyrus (medial) | R | 4.33 | 24 | 44 | 35 |
|  |  | Middle temporal gyrus | L | 4.33 | 44 | 25 | 21 |
|  |  | Rolandic operculum | R | 4.31 | 12 | 32 | 29 |
|  |  | Inferior temporal gyrus | L | 4.31 | 42 | 26 | 14 |
|  |  | Angular gyrus | L | 4.3 | 40 | 13 | 33 |
|  |  | Putamen | L | 4.23 | 34 | 31 | 20 |
|  |  | Insula | R | 4.14 | 13 | 32 | 29 |
|  |  | Hippocampus | R | 4.04 | 16 | 24 | 24 |
|  |  | Thalamus | R | 4.03 | 22 | 29 | 23 |
|  |  | Superior frontal gyrus (dorsal) | R | 3.97 | 20 | 45 | 40 |
|  |  | Inferior parietal lobule | R | 3.93 | 9 | 18 | 38 |
|  |  | Rolandic operculum | L | 3.93 | 37 | 32 | 28 |
|  |  | Supramarginal gyrus | R | 3.9 | 11 | 32 | 33 |
|  |  | Amygdala | L | 3.88 | 33 | 32 | 19 |
|  |  | Superior frontal gyrus (medial) | L | 3.88 | 25 | 49 | 36 |
|  |  | Supplementary motor area | R | 3.86 | 23 | 44 | 39 |
|  |  | Inferior occipital gyrus | L | 3.84 | 36 | 12 | 19 |
|  |  | Anterior cingulate gyrus | L | 3.83 | 25 | 46 | 27 |
|  |  | Insula | L | 3.81 | 37 | 31 | 28 |
|  |  | Inferior frontal gyrus (triangular) | L | 3.79 | 38 | 41 | 31 |
|  |  | Thalamus | L | 3.77 | 25 | 27 | 24 |
|  |  | Parahippocampal gyrus | L | 3.76 | 34 | 22 | 20 |
|  |  | Orbitofrontal cortex (superior) | R | 3.71 | 20 | 43 | 15 |
|  |  | Inferior frontal gyrus (opercular) | L | 3.71 | 37 | 37 | 32 |
|  |  | Orbitofrontal cortex (inferior) | R | 3.68 | 10 | 43 | 20 |
|  |  | Supramarginal gyrus | L | 3.66 | 45 | 32 | 35 |
|  |  | Precentral gyrus | L | 3.66 | 35 | 37 | 37 |
|  |  | Inferior parietal lobule | L | 3.63 | 35 | 14 | 37 |
|  |  | Parahippocampal gyrus | R | 3.63 | 17 | 27 | 16 |
|  |  | Rectus gyrus | L | 3.62 | 25 | 43 | 15 |
|  |  | Superior temporal gyrus | R | 3.61 | 6 | 21 | 33 |
|  |  | Supplementary motor area | L | 3.58 | 29 | 41 | 41 |
|  |  | Superior temporal gyrus | L | 3.53 | 37 | 27 | 24 |
|  |  | Anterior cingulate gyrus | R | 3.53 | 24 | 46 | 27 |
|  |  | Middle occipital gyrus | L | 3.48 | 40 | 10 | 31 |
|  |  | Caudate | R | 3.37 | 19 | 37 | 28 |
|  |  | Pallidum | L | 3.34 | 32 | 33 | 25 |
|  |  | Rectus gyrus | R | 3.28 | 23 | 41 | 16 |
|  |  | Olfactory cortex | L | 3.18 | 25 | 41 | 19 |
|  |  | Lingual gyrus | R | 3.14 | 22 | 12 | 21 |
|  |  | Inferior temporal gyrus | R | 3.11 | 5 | 19 | 19 |
|  | SocialCognitive (L) | Supplementary motor area | L | 8.53 | 25 | 30 | 38 |
|  |  | Middle cingulate gyrus | L | 8.25 | 25 | 30 | 37 |
|  |  | Supplementary motor area | R | 8.23 | 24 | 30 | 38 |
|  |  | Middle cingulate gyrus | R | 8.18 | 24 | 31 | 37 |
|  |  | Paracentral lobule | L | 7.77 | 25 | 29 | 38 |
|  |  | Thalamus | L | 7.76 | 29 | 28 | 24 |
|  |  | Thalamus | R | 7.25 | 20 | 28 | 23 |
|  |  | Paracentral lobule | R | 7.23 | 23 | 27 | 38 |
|  |  | Superior temporal gyrus | R | 7.19 | 4 | 30 | 28 |
|  |  | Superior temporal gyrus | L | 7 | 43 | 27 | 27 |
|  |  | Putamen | L | 6.97 | 34 | 29 | 25 |
|  |  | Rolandic operculum | L | 6.82 | 36 | 27 | 30 |
|  |  | Postcentral gyrus | L | 6.68 | 42 | 30 | 38 |
|  |  | Insula | R | 6.66 | 14 | 35 | 26 |
|  |  | Rolandic operculum | R | 6.52 | 12 | 28 | 30 |
|  |  | Precuneus | R | 6.38 | 24 | 25 | 40 |
|  |  | Putamen | R | 6.33 | 15 | 32 | 28 |
|  |  | Precuneus | L | 6.29 | 25 | 23 | 41 |
|  |  | Insula | L | 6.17 | 34 | 29 | 26 |
|  |  | Postcentral gyrus | R | 6.17 | 8 | 33 | 37 |
|  |  | Inferior parietal lobule | L | 6.13 | 43 | 30 | 37 |
|  |  | Heschl's gyrus | R | 6.07 | 12 | 31 | 28 |
|  |  | Heschl's gyrus | L | 6.04 | 38 | 30 | 25 |
|  |  | Precentral gyrus | L | 6.02 | 31 | 29 | 44 |
|  |  | Supramarginal gyrus | R | 5.94 | 3 | 31 | 30 |
|  |  | Cuneus | L | 5.87 | 30 | 16 | 29 |
|  |  | Precentral gyrus | R | 5.74 | 19 | 31 | 44 |
|  |  | Amygdala | L | 5.68 | 31 | 32 | 20 |
|  |  | Fusiform gyrus | R | 5.42 | 18 | 24 | 18 |
|  |  | Calcarine cortex | R | 5.39 | 18 | 18 | 24 |
|  |  | Supramarginal gyrus | L | 5.35 | 46 | 30 | 27 |
|  |  | Lingual gyrus | R | 5.27 | 20 | 27 | 21 |
|  |  | Pallidum | L | 5.27 | 31 | 31 | 22 |
|  |  | Lingual gyrus | L | 5.27 | 31 | 20 | 21 |
|  |  | Calcarine cortex | L | 5.15 | 29 | 20 | 23 |
|  |  | Cuneus | R | 5.13 | 19 | 12 | 31 |
|  |  | Superior parietal gyrus | L | 5.08 | 31 | 25 | 43 |
|  |  | Superior frontal gyrus (dorsal) | R | 5.04 | 20 | 32 | 44 |
|  |  | Parahippocampal gyrus | R | 4.73 | 18 | 25 | 18 |
|  |  | Rectus gyrus | R | 4.73 | 23 | 47 | 15 |
|  |  | Superior parietal gyrus | R | 4.71 | 21 | 24 | 43 |
|  |  | Pallidum | R | 4.66 | 18 | 32 | 22 |
|  |  | Hippocampus | L | 4.63 | 31 | 31 | 15 |
|  |  | Superior frontal gyrus (dorsal) | L | 4.61 | 30 | 32 | 45 |
|  |  | Superior occipital gyrus | L | 4.54 | 31 | 14 | 30 |
|  |  | Middle temporal gyrus | R | 4.42 | 3 | 26 | 25 |
|  |  | Middle temporal gyrus | L | 4.37 | 45 | 21 | 26 |
|  |  | Temporal pole (superior) | R | 4.3 | 5 | 39 | 20 |
|  |  | Fusiform gyrus | L | 4.26 | 32 | 21 | 19 |
|  |  | Parahippocampal gyrus | L | 4.11 | 29 | 23 | 19 |
|  |  | Superior occipital gyrus | R | 3.77 | 18 | 12 | 34 |
|  |  | Orbitofrontal cortex (medial) | R | 3.66 | 24 | 48 | 17 |
|  |  | Rectus gyrus | L | 3.57 | 25 | 47 | 15 |
|  |  | Orbitofrontal cortex (medial) | L | 3.53 | 26 | 49 | 18 |
|  |  | Temporal pole (superior) | L | 3.46 | 40 | 37 | 19 |
|  |  | Hippocampus | R | 3.33 | 20 | 30 | 19 |
|  |  | Posterior cingulate gyrus | R | 3.29 | 22 | 22 | 23 |
|  |  | Inferior parietal lobule | R | 3.11 | 13 | 27 | 39 |
|  | SocialCognitive (R) | Middle cingulate gyrus | R | 7.93 | 24 | 31 | 37 |
|  |  | Thalamus | R | 7.85 | 19 | 29 | 25 |
|  |  | Middle cingulate gyrus | L | 7.71 | 25 | 30 | 37 |
|  |  | Supplementary motor area | R | 7.7 | 24 | 31 | 38 |
|  |  | Supplementary motor area | L | 7.42 | 25 | 30 | 38 |
|  |  | Paracentral lobule | R | 7.09 | 23 | 27 | 38 |
|  |  | Paracentral lobule | L | 7.07 | 25 | 29 | 38 |
|  |  | Putamen | L | 6.85 | 33 | 32 | 25 |
|  |  | Thalamus | L | 6.75 | 29 | 28 | 24 |
|  |  | Precentral gyrus | R | 6.71 | 18 | 32 | 44 |
|  |  | Superior temporal gyrus | L | 6.58 | 39 | 26 | 28 |
|  |  | Rolandic operculum | L | 6.57 | 38 | 26 | 29 |
|  |  | Superior temporal gyrus | R | 6.56 | 8 | 30 | 28 |
|  |  | Precentral gyrus | L | 6.51 | 30 | 30 | 44 |
|  |  | Rolandic operculum | R | 6.5 | 11 | 32 | 30 |
|  |  | Pallidum | L | 6.3 | 32 | 32 | 25 |
|  |  | Precuneus | L | 6.28 | 31 | 20 | 23 |
|  |  | Pallidum | R | 6.19 | 18 | 31 | 22 |
|  |  | Postcentral gyrus | L | 6.17 | 39 | 32 | 40 |
|  |  | Insula | R | 6.16 | 14 | 29 | 26 |
|  |  | Calcarine cortex | L | 6.14 | 31 | 18 | 24 |
|  |  | Lingual gyrus | L | 6.04 | 31 | 20 | 22 |
|  |  | Putamen | R | 6.02 | 14 | 35 | 24 |
|  |  | Lingual gyrus | R | 5.98 | 20 | 27 | 21 |
|  |  | Postcentral gyrus | R | 5.93 | 21 | 26 | 42 |
|  |  | Insula | L | 5.92 | 34 | 27 | 30 |
|  |  | Superior frontal gyrus (dorsal) | R | 5.86 | 18 | 33 | 44 |
|  |  | Heschl's gyrus | R | 5.75 | 9 | 31 | 27 |
|  |  | Precuneus | R | 5.71 | 23 | 23 | 42 |
|  |  | Calcarine cortex | R | 5.61 | 20 | 18 | 25 |
|  |  | Cuneus | L | 5.46 | 28 | 11 | 33 |
|  |  | Fusiform gyrus | R | 5.13 | 16 | 21 | 20 |
|  |  | Supramarginal gyrus | R | 5.09 | 11 | 32 | 33 |
|  |  | Inferior parietal lobule | L | 5.01 | 43 | 30 | 37 |
|  |  | Cuneus | R | 4.96 | 21 | 17 | 29 |
|  |  | Heschl's gyrus | L | 4.96 | 39 | 30 | 27 |
|  |  | Superior parietal gyrus | R | 4.83 | 20 | 24 | 42 |
|  |  | Superior parietal gyrus | L | 4.8 | 31 | 25 | 43 |
|  |  | Inferior parietal lobule | R | 4.8 | 13 | 27 | 39 |
|  |  | Hippocampus | R | 4.79 | 16 | 30 | 16 |
|  |  | Parahippocampal gyrus | R | 4.76 | 17 | 28 | 16 |
|  |  | Hippocampus | L | 4.74 | 29 | 26 | 21 |
|  |  | Supramarginal gyrus | L | 4.68 | 44 | 30 | 35 |
|  |  | Superior occipital gyrus | L | 4.67 | 31 | 14 | 30 |
|  |  | Superior frontal gyrus (dorsal) | L | 4.67 | 30 | 32 | 45 |
|  |  | Rectus gyrus | R | 4.51 | 23 | 48 | 15 |
|  |  | Orbitofrontal cortex (medial) | R | 4.34 | 24 | 44 | 18 |
|  |  | Amygdala | L | 4.34 | 30 | 32 | 20 |
|  |  | Temporal pole (superior) | R | 4.16 | 8 | 37 | 23 |
|  |  | Posterior cingulate gyrus | L | 4.07 | 26 | 21 | 23 |
|  |  | Posterior cingulate gyrus | R | 4.05 | 22 | 22 | 23 |
|  |  | Middle frontal gyrus | R | 3.92 | 10 | 36 | 40 |
|  |  | Superior occipital gyrus | R | 3.83 | 19 | 8 | 34 |
|  |  | Fusiform gyrus | L | 3.83 | 30 | 22 | 18 |
|  |  | Parahippocampal gyrus | L | 3.77 | 28 | 25 | 18 |
|  |  | Anterior cingulate gyrus | L | 3.64 | 25 | 43 | 19 |
|  |  | Rectus gyrus | L | 3.53 | 24 | 45 | 16 |
|  |  | Middle temporal gyrus | R | 3.5 | 3 | 26 | 25 |
|  |  | Middle occipital gyrus | R | 3.43 | 16 | 11 | 33 |
|  |  | Orbitofrontal cortex (superior) | R | 3.24 | 21 | 48 | 14 |
